# Development and internal validation of a prediction model for sleep apnea syndrome treated with continuous positive airway pressure based on claims and health checkup data linked to personal health records

**DOI:** 10.64898/2026.02.08.26345272

**Authors:** Tatsuya Muraki, Tsuyoshi Ueda, Chihiro Hasegawa, Hiroshi Usui, Hiroshi Koshimizu, Kenichi Ariyada, Kunio Kusajima, Yasuhiro Tomita, Masashi Yanagisawa, Masao Iwagami

## Abstract

**Purpose:** To develop and validate a prediction model for sleep apnea syndrome (SAS) treated with continuous positive airway pressure (CPAP) in the general population.

**Methods:** Using claims and health checkup data held by JMDC Inc., linked to personal health records (Pep Up), we developed and internally validated a prediction model for SAS treated with CPAP, defined as a diagnosis of SAS and reimbursement records of CPAP. Every three months from January 1, 2022 to July 1, 2024 (i.e., 11 timepoints), we identified eligible individuals with available data both 1 year before and 1 year after that timepoint to define the presence/absence of SAS treated with CPAP, as well as 279 predictor variables. We developed a LightGBM model for the training and tuning datasets and evaluated its performance on the validation dataset.

**Results:** Among 18,692,873 observations (mean age 44.8±11.3 years, women 37.5%) obtained from 1,858,566 people, 300,868 (1.6%) had SAS treated with CPAP. The area under the receiver operating characteristic curve was 0.898 (95% confidence interval 0.895–0.901). The positive predictive values among people with the top 1% and 10% prediction scores were 28.3% and 10.3%, respectively. According to the SHapley Additive exPlanations plot, male sex was the most important predictor, followed by age, body mass index, and waist circumference. We also demonstrated that personal health records significantly improved the predictive performance.

**Conclusion:** We developed a prediction model to identify people at high risk of SAS and encourage them to undergo polysomnography or related tests.

## Introduction

Sleep apnea syndrome (SAS), mainly obstructive sleep apnea (OSA), is a common disease [1–3], and is associated with a variety of morbidities and mortality [4–6]. Early detection and intervention for SAS are important for improving patient prognosis [7–9]. However, many people with SAS remain undiagnosed in the community [10–13]. This may be because many people with SAS find it difficult to recognize the condition themselves. Even if they have symptoms such as daytime sleepiness, their capacity for and access to polysomnography (PSG) are limited in most countries, including Japan, preventing timely diagnosis and treatment such as continuous positive airway pressure (CPAP) [13–15].

Therefore, identifying or predicting people at high risk of SAS from any available health information is warranted to encourage them to undergo PSG or related tests, such as a pulse oximeter and an airflow sensor at home [16] or portable encephalography [17]. Many studies have developed and validated diagnostic prediction models for SAS or OSA based on electronic health records, claims data, and/or personal health records (PHRs) obtained from mobile apps and wearable or home medical devices [18–21].

However, in most previous studies, the study population consisted of people receiving PSG or related tests [18–21]. Consequently, the sample size was often small. Moreover, the study populations did not represent the general population, and the established models cannot be used to suggest who should be prioritized for PSG or related tests. For a prediction model for SAS to be widely used in the community, the study population should be broadly representative. In addition, predictor variables should ideally be obtained from routinely collected health data, such as medical claims and PHRs.

In Japan, JMDC Inc. has collected medical claims data and health checkup data of people with employee insurance since 2005, and PHRs from mobile apps and wearable or home medical devices have recently been linked from agreeing members within the system of “Pep Up” [22, 23]. Using these data, we aimed to develop and internally validate a prediction model for SAS treated with CPAP.

## Methods

### Data source

This study utilized data obtained from Pep Up [22, 23], an information and communications technology service provided by JMDC Inc. for society-managed health insurance members, on top of the JMDC payer database [24]. As of July 2025, there were approximately two million Pep Up users.

Informed consent was obtained from individuals in the Pep Up system to use their lifelog data (as detailed below), claims data, and annual health checkup results in the JMDC payer database for research. The data were pseudonymized and processed in pseudonymized form by JMDC Inc. This study was conducted in accordance with the ethical standards of the Declaration of Helsinki and was approved by the Ethics Committee of the Research Institute of Healthcare Data Science (approval number: RI2025004).

### Study design and population

This study aimed to establish a “diagnostic” prediction model for presence/absence of SAS treated with CPAP cross-sectionally, in contrast to a “prognostic” prediction model for the future incidence of SAS [25]. However, because of the nature of routinely collected real-world data and/or PHRs, a certain span (instead of a single day) is required for health information to be recorded and used to define the presence/absence of outcome and predictor variables. Therefore, we first determined the *index* dates for prediction, such as January 1, 2022 (**Figure 1**). We used information recorded both 1 year before and 1 year after the index date to define the outcome. Thus, for this index date, we identified and included people registered with Pep Up during the whole 2 years. We repeated this process every three months from January 1, 2022 to July 1, 2024 (i.e., 11 timepoints) to create the analysis dataset. Thus, each person contributed to the analysis dataset from once at minimum up to a maximum of 11 times.

**Figure 1.**
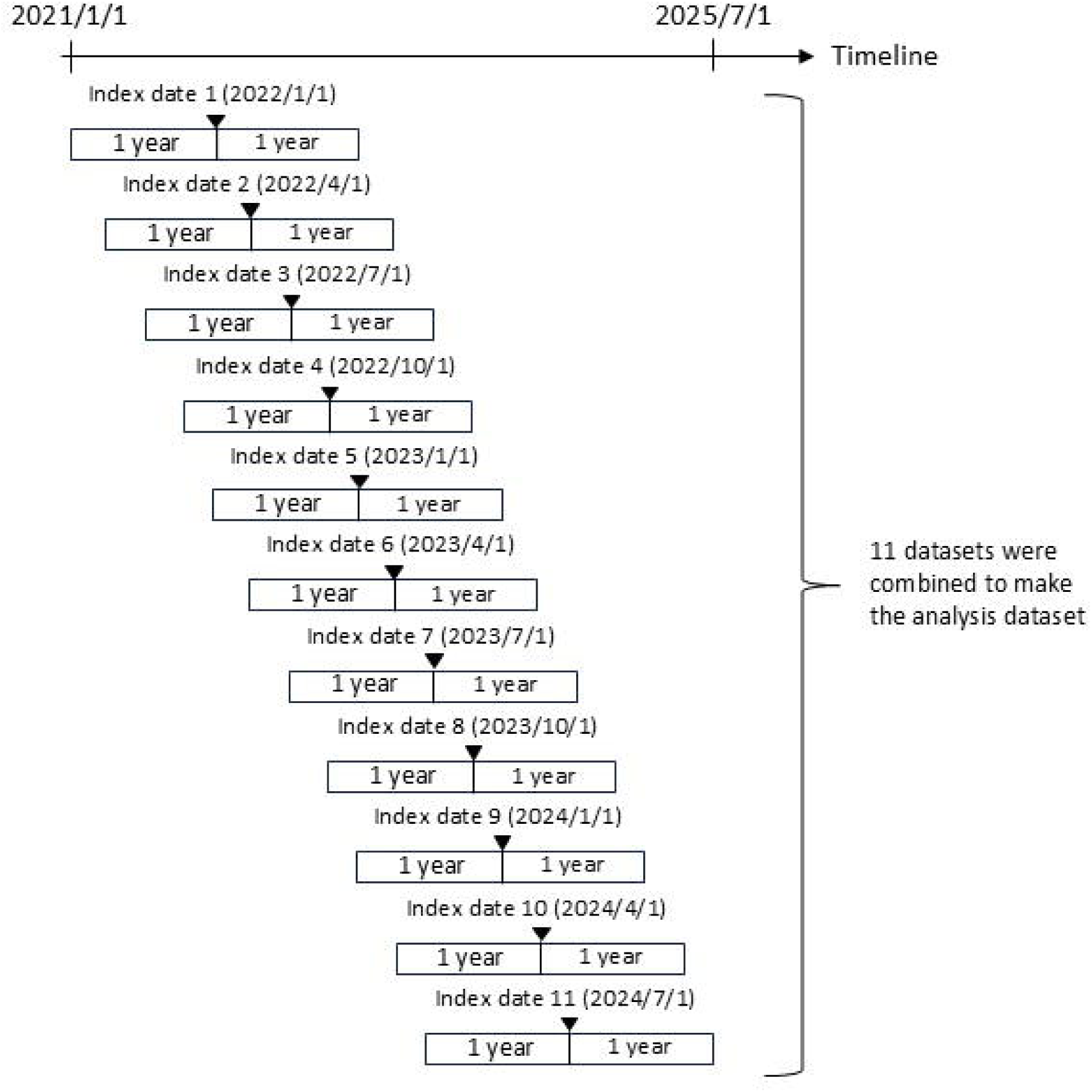
Graphical representation of creating the analysis dataset. Every 3 months from January 1, 2022 to July 1, 2024 (i.e., 11 timepoints), we identified and included people registered in Pep Up during the entire 2 years, both 1 year before and 1 year after the index date, to define the outcome (i.e., sleep apnea syndrome treated with continuous positive airway pressure)

### Outcome

The outcome was SAS treated with CPAP, defined as a diagnosis (excluding “suspected” diagnoses) of SAS, recorded as Japanese diagnostic codes corresponding to International Classification of Disease-10 code G47.3, in addition to CPAP, recorded as Japanese procedure codes C107-2, C171-2, and C165 in Japanese insurance claims, during the 2 years. Notably, in Japan, CPAP for SAS is covered by health insurance and is reimbursed if the apnea–hypopnea index (AHI) is 20 or higher by PSG or if the AHI is 40 or higher by a simple test, such as a pulse oximeter and an airflow sensor at home [26]. The government constantly monitors whether an indication is appropriate to prevent fraudulent or false claims, meaning that the specificity of CPAP for SAS is expected to be extremely high.

### Predictor variables

From the claims data and annual health checkup results in the JMDC payer database and the PHRs in Pep Up, we extracted information on 279 predictor variables (**Online Resource 1**). The predictor variables were as follows: sex and age at the index date; 50 disease diagnoses in the medical claims data, for which the number of months during 1 year before the index month (not including the index month itself) was counted, providing a number from 0–12; 94 types of drug (corresponding to World Health Organization Anatomical Therapeutic Chemical Classification 3-digit codes) prescriptions in the medical claims data, for which the number of months during 1 year before the index month (not including the index month itself) was counted, providing a number from 0–12; health checkup results (23 variables) at the closest day before the index date during the 2 years before the index date; lifelog data (110 variables) recorded in Pep Up, linked to devices including home blood-pressure monitors, smartwatches, and body composition monitors, or manual inputs by the users, from 90 days to 1 day before the index date.

### Statistical analysis

After summarizing the distribution of the predictor and outcome variables in the analysis dataset, we randomly split the study participants (regardless of the number of contributions to the analysis dataset) into training, tuning, and validation datasets in a ratio of 6:2:2. The split was performed at the person level, meaning that observations (records) from the same person belonged only to the training, tuning, and validation datasets. We then developed a prediction model in the training and tuning datasets and assessed its predictive performance in the validation dataset.

The model was created using LightGBM, a type of gradient-boosting decision tree, owing to its high predictive performance and computational efficiency [27]. We used log-loss as the loss function. Hyperparameters were determined via a grid search to minimize log loss in the tuning data. The search ranges were set as num_leaves = 32, 128, and 512; min_child_samples = 10, 100, 1,000, and 10,000; and feature_fraction = 0.6, 0.8, and 1.0. The n_estimators were determined by early stopping. To ensure clinical interpretability, we applied monotonic constraints to the predictor variables (**Online Resource 1**). Missing values were input into the LightGBM model because it could handle missing data. In tree-based algorithms, the model learns the optimal branch directions to handle missing values during training.

As a metric of model performance, we estimated the area under the receiver operating characteristic curve (AUROC). In addition, we estimated the positive predictive value (PPV), which is the proportion of individuals with SAS treated with CPAP (i.e., true cases) among individuals with the top 1% and 10% of the prediction score, respectively. Finally, the SHapley Additive exPlanations (SHAP) function was used to interpret the model via a SHAP plot (beeswarm plot) of the top 20 predictor variables [28]. This plot visualizes the distribution of the SHAP values for each feature, demonstrating both the magnitude and direction of their contribution to the model output on the log scale.

A subgroup analysis was conducted based on sex. As a sensitivity analysis, we first assessed the predictive ability of the model in the validation datasets split at 11 timepoints (**Figure 1**) separately to confirm that prediction ability is not affected by year or season. Second, we developed and validated a prognostic prediction model (in contrast to a diagnostic prediction model in our main analysis) for the new incidence of SAS treated with CPAP within 1 year after the index date, by excluding patients with any SAS diagnosis (i.e., both confirmed and “suspected” SAS diagnosis, with or without CPAP) during the past year before the index date.

As an additional analysis, to quantify the contribution of PHRs in Pep Up (110 variables) to the prediction, we compared the original model based on all 279 predictor variables and the model without PHRs in Pep Up in terms of AUROC. We also estimated the integrated discrimination improvement (IDI), which is the improvement in the differences between the average predicted probabilities of cases and non-cases [29]. For this purpose, we restricted the study population to (i) people with at least one non-missing variable (among 110 variables) in PHRs, suggesting that they are “light users” of Pep Up, and (ii) people with ≥30-day recordings (among 90 days before the index date) of systolic blood pressure, diastolic blood pressure, resting heart rate, body weight, body fat percentage, steps, and sleep duration, suggesting that they are “heavy users” of Pep Up.

All data management and statistical analyses were performed using Python (version 3.12.9).

## Results

From January 1, 2022 to July 1, 2024, 1,858,566 people contributed to the analysis dataset at least once (up to 11 times), totaling to 18,692,873 observations (mean age 44.8±11.3 years, women 37.5%). The distribution of the 279 predictor variables is shown in **Online Resource 2**.

Among the 18,692,873 observations (records), 300,868 (1.6%) had SAS treated with CPAP. The prevalences according to sex, age, and BMI are shown in **Figure 2**. The AUROC was estimated to be 0.898 (95% confidence interval [CI] 0.895–0.901; **Figure 3**). The PPVs among the participants with the top 1% and 10% prediction scores were 28.3% and 10.3%, respectively. According to the SHAP plot, male sex was the most important predictor, followed by age, BMI, and waist circumference (**Figure 4**).

**Figure 2.**
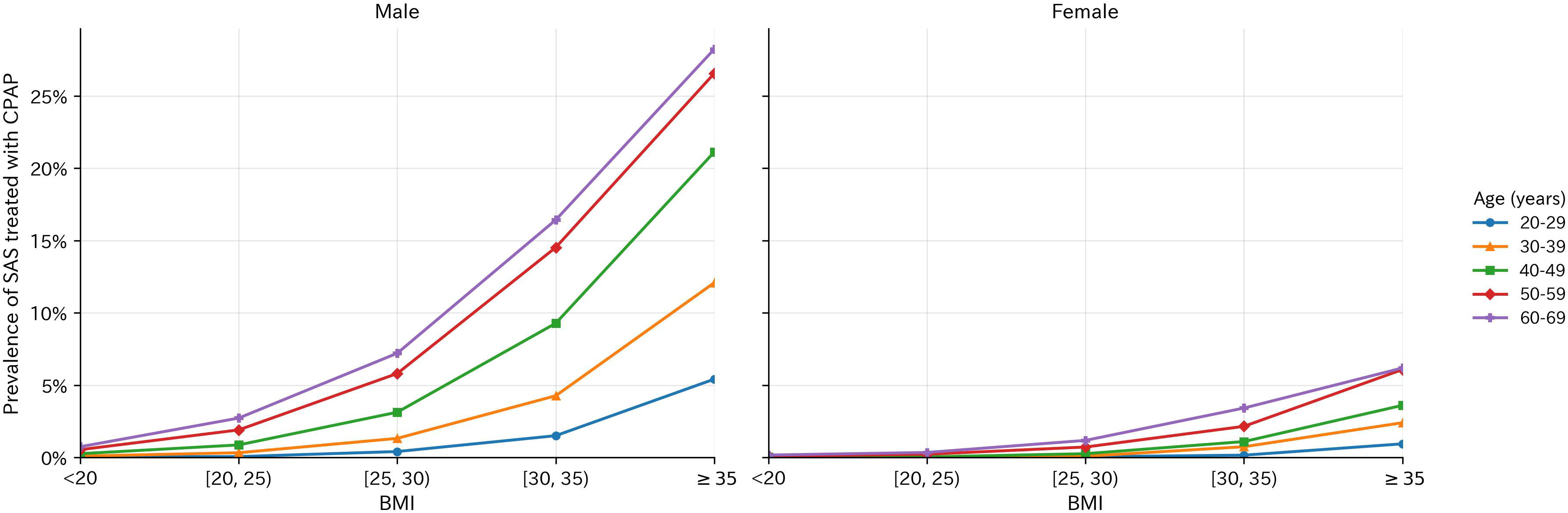
Prevalence of sleep apnea syndrome treated with continuous positive airway pressure according to sex, age, and body mass index. SAS, sleep apnea syndrome; CPAP, continuous positive airway pressure; BMI, body mass index

**Figure 3.**
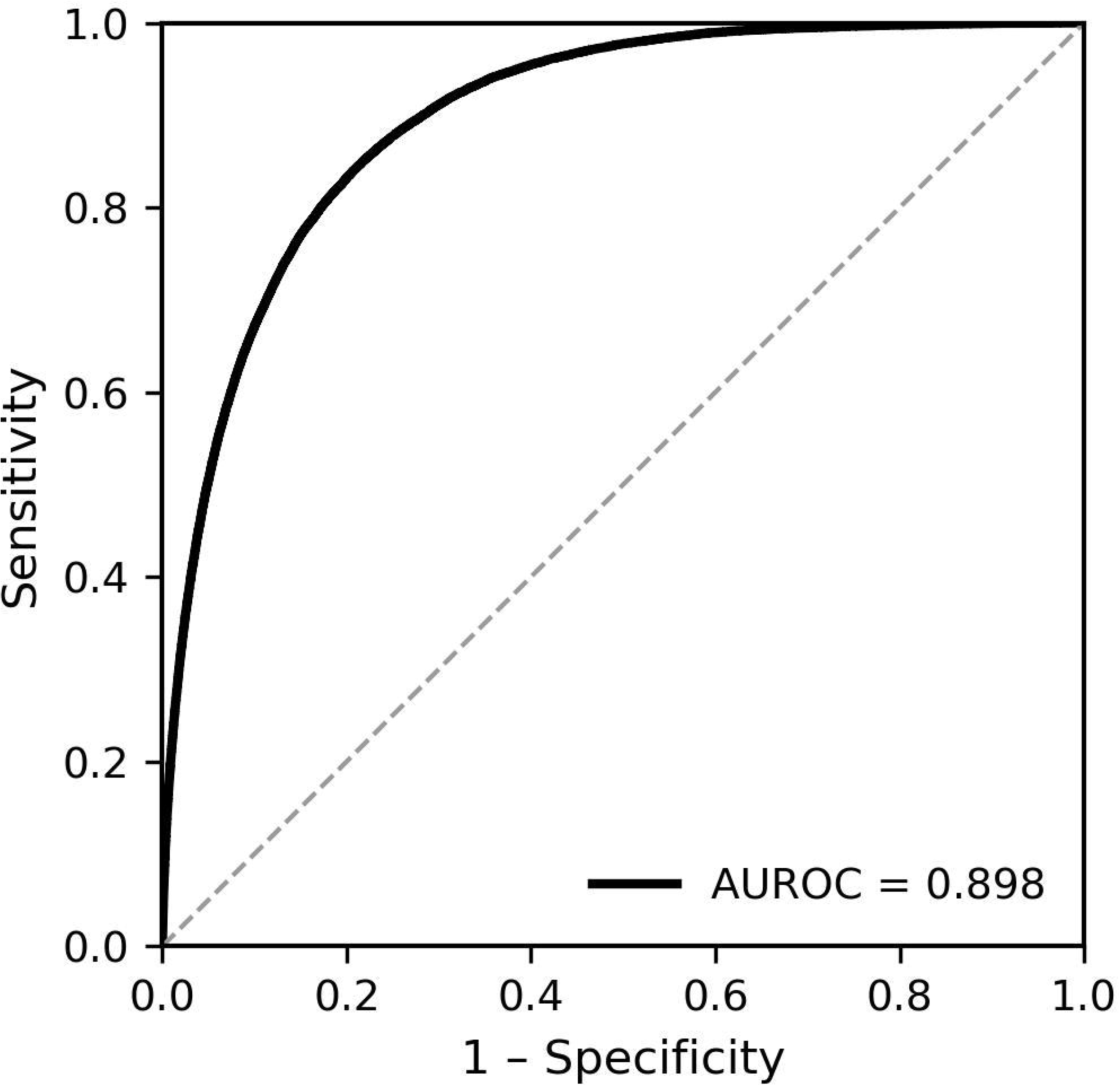
The receiver operating characteristic curve of the prediction model for sleep apnea syndrome treated with continuous positive airway pressure. AUROC, area under the receiver operating characteristic curve

**Figure 4.**
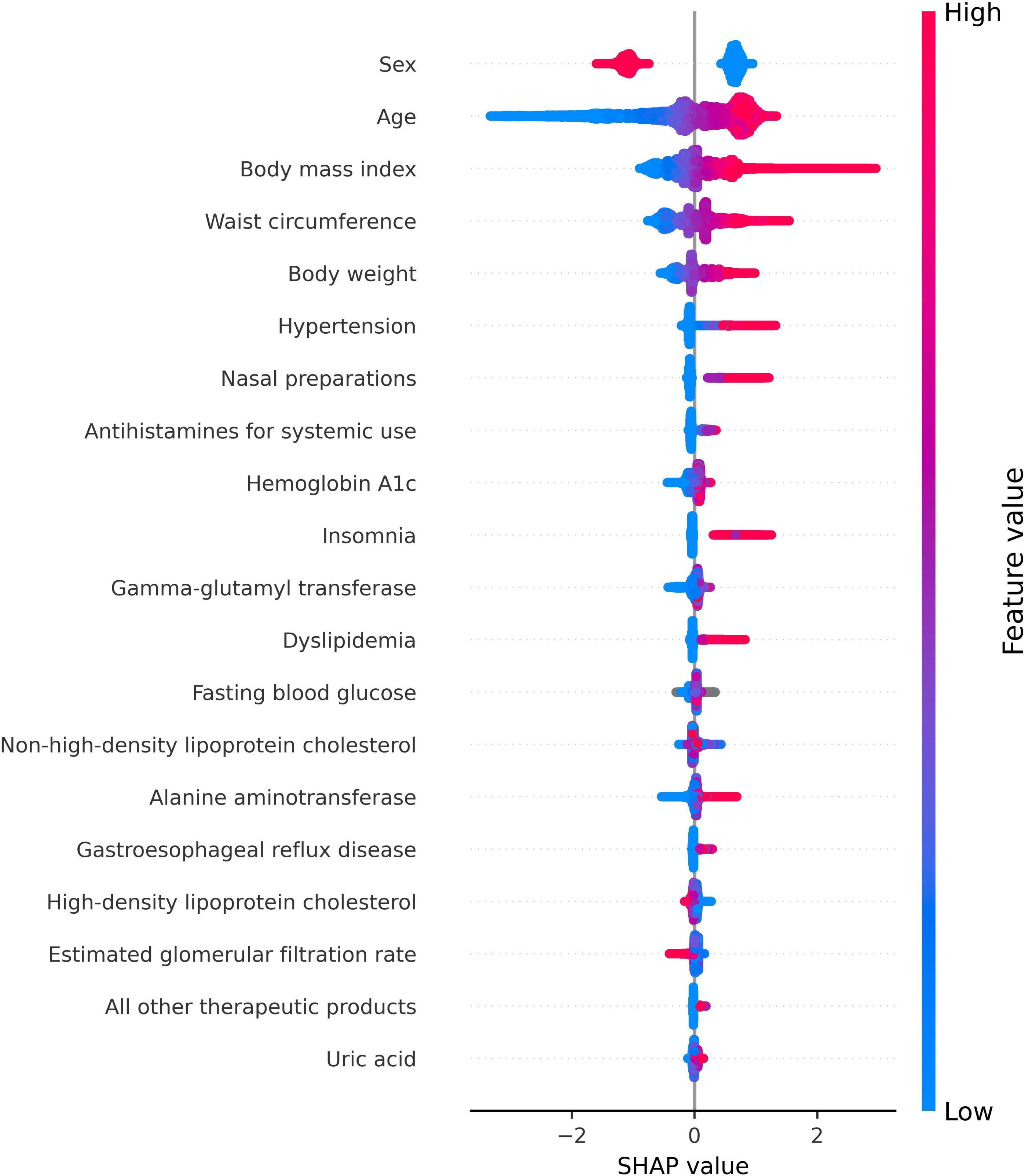
SHapley Additive exPlanations (SHAP) plot for the top 20 predictor variables in the prediction model for sleep apnea syndrome treated with continuous positive airway pressure.

In the subgroup analysis, the prevalences of SAS treated with CPAP were 2.4% and 0.2% in men and women, respectively. The AUROCs were 0.860 (95% CI 0.856–0.864) and 0.906 (95% CI 0.890–0.922) in men and women, respectively. The PPVs among people with the top 1% and 10% prediction scores were 32.7% and 13.3% in men and 7.2% and 1.7% in women, respectively. The SHAP plots are shown in **Online Resource 3.**

In the first sensitivity analysis, separate validation datasets at 11 timepoints yielded almost the same AUROC, suggesting that prediction ability was not affected by year or season (**Online Resource 4**). In the second sensitivity analysis to develop a prognostic prediction model for the new incidence of SAS treated with CPAP, among 18,326,135 observations (records) without any SAS diagnosis during the year before the index date, 39,906 (0.22%) had new SAS treated with CPAP during 1 year after the index date. The AUROC was 0.845 (95% CI 0.838–0.851).

Among 2,300,547 observations (records) obtained from 460,632 “light users” of Pep Up, 42,076 (1.8%) had SAS treated with CPAP. The AUROCs were 0.896 (95% CI 0.888–0.903) and 0.895 (95% CI 0.887–0.902) in the models with and without PHRs in Pep Up, respectively, indicating an increase in the AUROC (0.0012, 95% CI 0.0003–0.0021) with the use of PHRs in Pep Up. The IDI was 1.07% (95% CI 0.93–1.22), implying that the prediction model improved by 1.07% using PHRs in Pep Up. The top 20 ranking of the SHAP plot was generally similar to that of the original model in the main analysis, but the mean sleep duration during the three months recorded in Pep Up ranked at 14^th^ (**Figure 5**).

**Figure 5.**
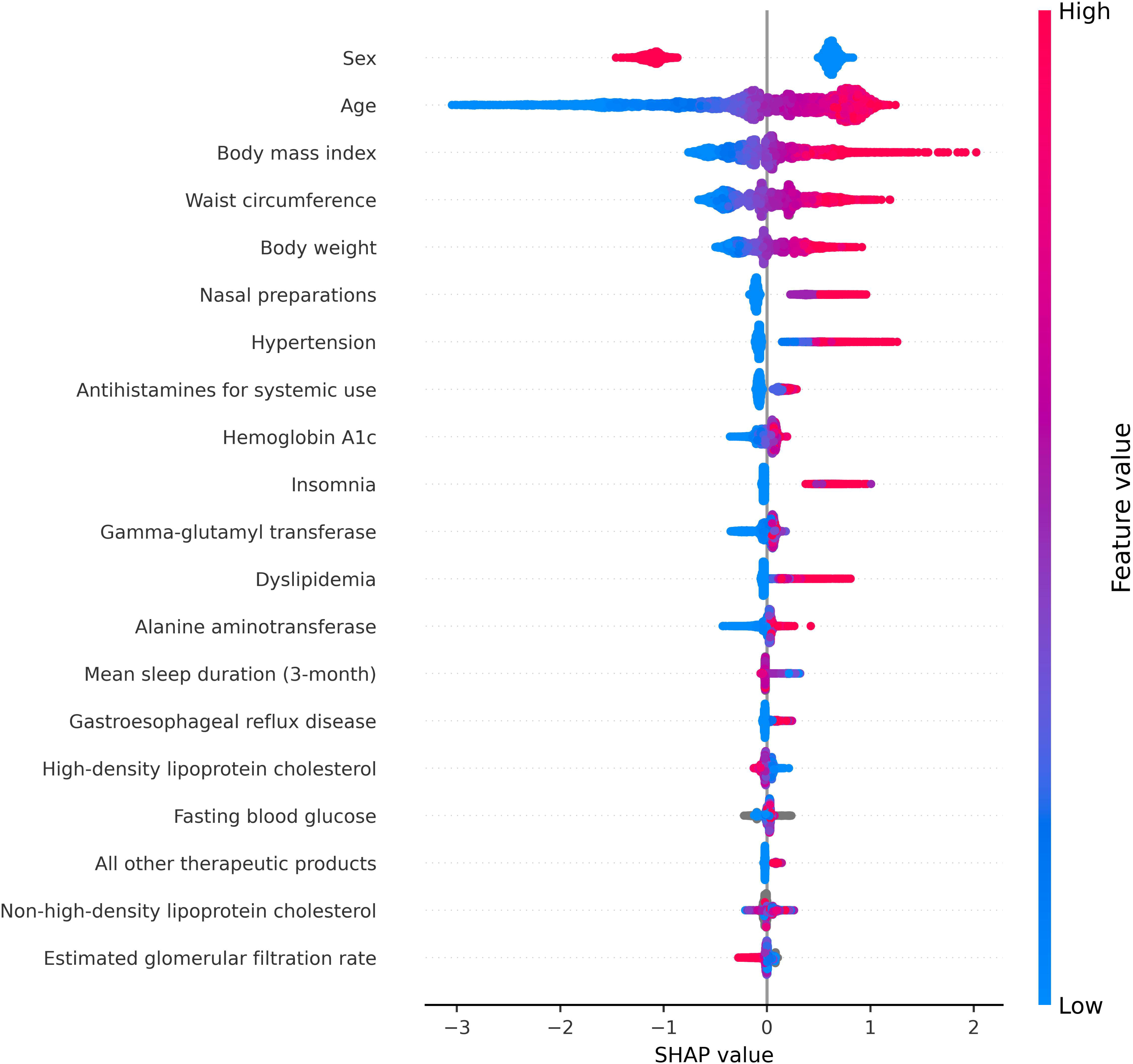
SHapley Additive exPlanations (SHAP) plot for the top 20 predictor variables in the prediction model for sleep apnea syndrome treated with continuous positive airway pressure among “light users” of Pep Up. “Light users” of Pep Up were defined as those with at least one non-missing variable among the 110 variables in their personal health records.

Among the 8,865 observations (records) obtained from 2,441 “heavy users” of Pep Up, 491 (5.5%) had SAS treated with CPAP. The AUROCs were 0.836 (95% CI 0.684–0.938) and 0.826 (95% CI 0.670–0.935) in the models with and without PHRs, respectively, indicating an increase in the AUROC (0.0096, 95% CI -0.0037–0.0239). The IDI was 4.57% (95% CI 2.22–6.67), implying that the prediction model improved by 4.57% using PHRs in Pep Up. Regarding the SHAP plot, five predictor variables based on PHRs in Pep Up were ranked between 9^th^ and 17^th^ (**Figure 6**).

**Figure 6.**
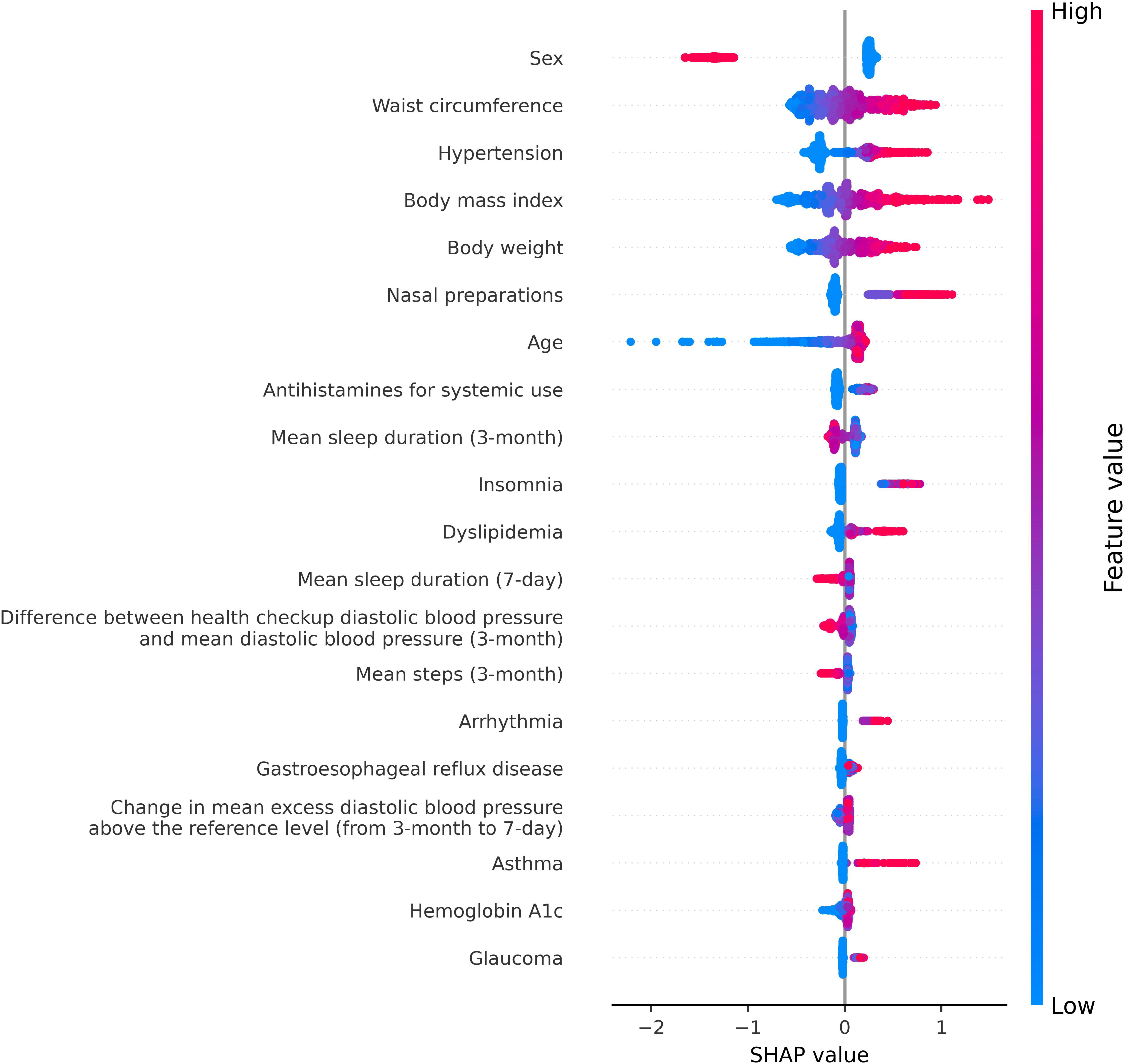
SHapley Additive exPlanations (SHAP) plot for the top 20 predictor variables in the prediction model for sleep apnea syndrome treated with continuous positive airway pressure among “heavy users” of Pep Up. “Heavy users” of Pep Up were defined as people with ≥30-day recordings (among 90 days before the index date) of systolic blood pressure, diastolic blood pressure, resting heart rate, body weight, body fat percentage, steps, and sleep duration.

## Discussion

Using claims and health checkup data linked to PHRs from mobile apps and wearable or home medical devices, we developed and internally validated a prediction model for SAS treated with CPAP. The model demonstrated a high predictive performance, with an AUROC of 0.898. Although the prevalence of SAS treated with CPAP was 1.6% in the study population, the PPVs among people with the top 1% and 10% of the prediction score were 28.3% and 10.3%, respectively, indicating that approximately 3 and 1 in 10 people would have SAS treated with CPAP, respectively. Considering the limited availability and high cost of inpatient PSG in many countries, including Japan [13–15], it seems efficient to encourage people with relatively high prediction scores to undergo PSG or related tests, such as a pulse oximeter and an airflow sensor at home [16] or portable encephalography [17].

Identifying patients with SAS through routine clinical practice or health checkups remains challenging. OSA is frequently underrecognized in primary care, largely because its cardinal symptoms, such as fatigue, insomnia, and uncontrolled hypertension, are non-specific and easily attributed to coexisting chronic conditions. Even patients who regularly visit primary care clinics may remain undiagnosed despite exhibiting a high pre-test probability of OSA [30]. In this context, a population-level prediction model that leverages routinely collected health data can provide valuable support for early diagnosis and timely referral within primary care workflows.

Well-known risk factors for SAS, including male sex, older age, higher body mass index, and higher waist circumference [18–21], were good predictors in our model. This study also quantified the contribution of PHRs to prediction in addition to claims data and health checkup results. Among “light users” of Pep Up, the improvements in AUROC and IDI were not large, though they were statistically significant. Only mean sleep duration during the three months was ranked among the top 20 predictor variables in terms of SHAP values. However, among “heavy users” of Pep Up, the improvements in AUROC and IDI became larger, and five predictor variables obtained from PHRs ranked. This indicates that the contribution of PHRs to the prediction of SAS treated with CPAP can be larger if users of PHR services use them more heavily and consistently. PHRs are a relatively new service, and the number of both light and heavy users of Pep Up has gradually increased. Further assessment is warranted after more PHRs accumulate in the Pep Up system.

More than 80% of individuals with moderate-to-severe OSA in the community are estimated to remain undiagnosed [31], underscoring the limitations of relying solely on sleep specialty clinics for case detection. A key strength of the present model is its ability to stratify risk using only health checkups and claims data, without imposing additional testing burdens. This makes it particularly suitable for large-scale implementation in corporate and municipal health programs. Moreover, integrating PHR data enables the model to extend its utility to individuals with limited access to outpatient care, including those receiving home-based medical services. This enhances opportunities to remotely identify people with potential OSA in underserved populations.

This study had several limitations. First, the outcome of the study was determining people with SAS treated with CPAP, which is the most severe form of SAS. Indeed, the prevalence of SAS treated with CPAP in the present study was only 1.6% (2.4% in men and 0.2% in women), lower than the expected prevalence of any type of SAS in the community: for example, the prevalence of SAS defined as AHI≥15 is estimated to be around 9.0% in men and 2.8% in women [32]. The present study indicates that only a small proportion of individuals with SAS reach CPAP treatment, particularly women. It is highly possible that some individuals without SAS treated with CPAP in the study population had undiagnosed or untreated SAS. Thus, a different study may be needed to develop a predictive model for any SAS (including undiagnosed/untreated SAS), although we expect that risk factors (and therefore, predictive models) would overlap between severe SAS and any SAS (including undiagnosed/untreated SAS). Second, despite being drawn from the general population, Pep Up users mainly consist of company employees and their family members and may be more health-conscious than non-users or the average Japanese population. Thus, the generalizability of these findings to other populations in Japan remains unknown. Similarly, the applicability of the study’s findings to populations in other countries should be carefully examined. Finally, there are many missing values for lifelog data among the overall studied population because not every Pep Up user has used devices, including home blood-pressure monitors, smartwatches, and body composition monitors, although the LightGBM model could deal with missing data without excluding individuals with missing values. PHRs significantly contributed to the prediction model among light and heavy users of Pep Up, yet it is expected that more lifelog data will accumulate in the Pep Up system.

OSA represents a substantial public health burden [31], and significant disparities in access to sleep medicine services have been well documented. A recent analysis in the United States estimated that approximately 45.8 million individuals reside in areas with limited access to sleep healthcare [33]. By integrating PHRs, health checkup data, and claims data, the proposed prediction model may mitigate such disparities by identifying high-risk individuals earlier before they reach specialized care. This has important implications not only for clinical practice, but also for community health initiatives, occupational health programs, and regional public health planning.

In conclusion, we developed a prediction model for SAS treated with CPAP based on claims data and health checkup data linked to PHRs, showing good predictive ability. This model could be useful to provide feedback and encourage people with high prediction scores to receive PSG or related tests for definitive diagnosis and treatment.

## Supporting information

Online Resource 1

Online Resource 2

Online Resource 3

Online Resource 4

## Author contributions

TM, TU, CH, and MI planned the study. TM, TU, CH, HU, and HK collected data. TM analyzed the data. TM and MI wrote the manuscript. TM prepared the tables, figures, and supplementary materials. KA, KK, YT, and MY critically commented on the first draft and contributed substantially to this study. All the authors reviewed the final version of the manuscript.

## Data availability statement

The datasets presented in this article are not readily available because we obtained data from JMDC Inc. and did not obtain permission to share these data with other parties. Researchers who meet the access criteria can acquire de-identified participant data from JMDC Inc. (https://www.jmdc.co.jp/en/). Requests to access the datasets should be directed to https://www.jmdc.co.jp/en/.

