## Supplementary material for "Development and internal validation of a prediction model for sleep apnea syndrome treated with continuous positive airway pressure based on claims and health checkup data linked to personal health records": Online Resource 1

**Online Resource 1. Definition of predictor variables**

|  | Variable name | Definition | Monotonic constraint |
| --- | --- | --- | --- |
| Basic information (2 variables) | | | |
| 1 | Sex | Information recorded in the JMDC payer database (encoded as 1 for men and 2 for women) | Negative |
| 2 | Age | Information recorded in the JMDC payer database | Positive |
| Diagnoses in the claims data (50 variables) | | | |
| 1 | Cancer | ICD-10 codes D00-D09, C00-C14, C15-C26, C30-C39, C40-C41, C43-C44, C45-C49, C50-C50, C51-C58, C60-C63, C64-C68, C69-C72, C73-C75, C76-C80, C81-C96, D45, D46, D471, D473, D474, D475 | Positive |
| 2 | Polyp | ICD-10 codes K228, K317, K620, K621, K635, K638, K644, K828, K928, N288, N289, N362, N840, N841, N842, N843 | Positive |
| 3 | Myocardial infarction | ICD-10 codes I21, I22 | Positive |
| 4 | Angina pectoris | ICD-10 codes I20 | Positive |
| 5 | Hypertension | ICD-10 codes I10-I15 | Positive |
| 6 | Arrhythmia | ICD-10 codes I44, I45, I47, I48, I49 | Positive |
| 7 | Stroke | ICD-10 codes I60, I61, I63 | Positive |
| 8 | Dementia | ICD-10 codes F00-F09, G30 | Positive |
| 9 | Schizophrenia | ICD-10 codes F20-F29 | Positive |
| 10 | Depressive disorder | ICD-10 codes F30-F39 | Positive |
| 11 | Neurotic disorder | ICD-10 codes F40-F48 | Positive |
| 12 | Eating disorder | ICD-10 codes F50 | Positive |
| 13 | Developmental disorder | ICD-10 codes F70-F79, F80-F89, F90-F98 | Positive |
| 14 | Insomnia | ICD-10 codes G470, G471, G472, G474, G478, G479 | Positive |
| 15 | Autonomic nervous system disorder | ICD-10 codes G90 | Positive |
| 16 | Epilepsy | ICD-10 codes G40, G41 | Positive |
| 17 | Asthma | ICD-10 codes J45, J46 | Positive |
| 18 | Tuberculosis | ICD-10 codes A15-A19 | Positive |
| 19 | Chronic obstructive pulmonary disease | ICD-10 codes J44 | Positive |
| 20 | Gastroesophageal reflux disease | ICD-10 codes K20, K21, K29, K30 | Positive |
| 21 | Gastric or duodenal ulcer | ICD-10 codes K22, K25, K26, K27, K28 | Positive |
| 22 | Inflammatory bowel disease | ICD-10 codes K50, K51 | Positive |
| 23 | Hepatitis | ICD-10 codes B15-B19, K73, K701 | Positive |
| 24 | Liver cirrhosis | ICD-10 codes K74, K703 | Positive |
| 25 | Fatty liver disease | ICD-10 codes K760, K769 | Positive |
| 26 | Pancreatitis | ICD-10 codes K85, K86 | Positive |
| 27 | Nephritis | ICD-10 codes N00-N08 | Positive |
| 28 | Renal failure | ICD-10 codes N18, N19 | Positive |
| 29 | Urolithiasis | ICD-10 codes N20-N23 | Positive |
| 30 | Cataract | ICD-10 codes H25, H26 | Positive |
| 31 | Glaucoma | ICD-10 codes H40-H42 | Positive |
| 32 | Meniere’s disease | ICD-10 codes H810 | Positive |
| 33 | Cervical dysplasia | ICD-10 codes N87 | Positive |
| 34 | Uterine fibroid | ICD-10 codes D25 | Positive |
| 35 | Ovarian cyst | ICD-10 codes D27 | Positive |
| 36 | Mastopathy | ICD-10 codes N60-N64 | Positive |
| 37 | Miscarriage / preterm delivery | ICD-10 codes O00-O08, O60, O200 | Positive |
| 38 | Cesarean section | ICD-10 codes O82, O842 | Positive |
| 39 | Endometriosis | ICD-10 codes N80 | Positive |
| 40 | Fracture/rupture | ICD-10 codes S02, S08, S12, S22, S28, S32, S38, S42, S48, S52, S58, S62, S68, S72, S78, S82, S88, S92, S98, T02, T05, T08, T10, S06, S07, S09, S14, S16, S17, S19, S24, S27, S29, S34, S36, S37, S39, S46, S47, S49, S56, S57, S59, S66, S67, S69, S76, S77, S79, S86, S87, S89, S96, S97, S99, T04, T06, T07, T09, T11, T13, T14, T116, T136, T142, T147 | Positive |
| 41 | Diabetes mellitus | ICD-10 codes E10-E14 | Positive |
| 42 | Thyroid disease | ICD-10 codes E00-E07 | Positive |
| 43 | Dyslipidemia | ICD-10 codes E78 | Positive |
| 44 | Rheumatoid arthritis | ICD-10 codes M05, M06 | Positive |
| 45 | Osteoarthritis | ICD-10 codes M15-M19 | Positive |
| 46 | Collagen disease | ICD-10 codes M30-M36 | Positive |
| 47 | Intervertebral disc herniation | ICD-10 codes M50, M51 | Positive |
| 48 | Premenstrual syndrome | ICD-10 codes N943 | Positive |
| 49 | Hyperuricemia | ICD-10 codes E79 | Positive |
| 50 | Gout | ICD-10 codes M10 | Positive |
| Prescriptions in the claims data (94 variables) | | | |
| 1 | Stomatological preparations | WHO-ATC code A01 | Positive |
| 2 | Drugs for acid related disorders | WHO-ATC code A02 | Positive |
| 3 | Drugs for functional gastrointestinal disorders | WHO-ATC code A03 | Positive |
| 4 | Antiemetics and antinauseants | WHO-ATC code A04 | Positive |
| 5 | Bile and liver therapy | WHO-ATC code A05 | Positive |
| 6 | Drugs for constipation | WHO-ATC code A06 | Positive |
| 7 | Antidiarrheals, intestinal antiinflammatory/antiinfective agents | WHO-ATC code A07 | Positive |
| 8 | Antiobesity preparations, excl. diet products | WHO-ATC code A08 | Positive |
| 9 | Digestives, incl. enzymes | WHO-ATC code A09 | Positive |
| 10 | Drugs used in diabetes | WHO-ATC code A10 | Positive |
| 11 | Vitamins | WHO-ATC code A11 | Positive |
| 12 | Mineral supplements | WHO-ATC code A12 | Positive |
| 13 | Tonics | WHO-ATC code A13 | Positive |
| 14 | Anabolic agents for systemic use | WHO-ATC code A14 | Positive |
| 15 | Appetite stimulants | WHO-ATC code A15 | Positive |
| 16 | Other alimentary tract and metabolism products | WHO-ATC code A16 | Positive |
| 17 | Antithrombotic agents | WHO-ATC code B01 | Positive |
| 18 | Antihemorrhagics | WHO-ATC code B02 | Positive |
| 19 | Antianemic preparations | WHO-ATC code B03 | Positive |
| 20 | Blood substitutes and perfusion solutions | WHO-ATC code B05 | Positive |
| 21 | Other hematological agents | WHO-ATC code B06 | Positive |
| 22 | Cardiac therapy | WHO-ATC code C01 | Positive |
| 23 | Antihypertensives | WHO-ATC code C02 | Positive |
| 24 | Diuretics | WHO-ATC code C03 | Positive |
| 25 | Peripheral vasodilators | WHO-ATC code C04 | Positive |
| 26 | Vasoprotectives | WHO-ATC code C05 | Positive |
| 27 | Beta blocking agents | WHO-ATC code C07 | Positive |
| 28 | Calcium channel blockers | WHO-ATC code C08 | Positive |
| 29 | Agents acting on the renin-angiotensin system | WHO-ATC code C09 | Positive |
| 30 | Lipid modifying agents | WHO-ATC code C10 | Positive |
| 31 | Antifungals for dermatological use | WHO-ATC code D01 | Positive |
| 32 | Emollients and protectives | WHO-ATC code D02 | Positive |
| 33 | Preparations for treatment of wounds and ulcers | WHO-ATC code D03 | Positive |
| 34 | Antipruritics, incl. antihistamines, anesthetics, etc. | WHO-ATC code D04 | Positive |
| 35 | Antipsoriatics | WHO-ATC code D05 | Positive |
| 36 | Antibiotics and chemotherapeutics for dermatological use | WHO-ATC code D06 | Positive |
| 37 | Corticosteroids, dermatological preparations | WHO-ATC code D07 | Positive |
| 38 | Antiseptics and disinfectants | WHO-ATC code D08 | Positive |
| 39 | Medicated dressings | WHO-ATC code D09 | Positive |
| 40 | Anti-acne preparations | WHO-ATC code D10 | Positive |
| 41 | Other dermatological preparations | WHO-ATC code D11 | Positive |
| 42 | Gynecological antiinfectives and antiseptics | WHO-ATC code G01 | Positive |
| 43 | Other gynecologicals | WHO-ATC code G02 | Positive |
| 44 | Sex hormones and modulators of the genital system | WHO-ATC code G03 | Positive |
| 45 | Urologicals | WHO-ATC code G04 | Positive |
| 46 | Pituitary and hypothalamic hormones and analogues | WHO-ATC code H01 | Positive |
| 47 | Corticosteroids for systemic use | WHO-ATC code H02 | Positive |
| 48 | Thyroid therapy | WHO-ATC code H03 | Positive |
| 49 | Pancreatic hormones | WHO-ATC code H04 | Positive |
| 50 | Calcium homeostasis | WHO-ATC code H05 | Positive |
| 51 | Antibacterials for systemic use | WHO-ATC code J01 | Positive |
| 52 | Antimycotics for systemic use | WHO-ATC code J02 | Positive |
| 53 | Antimycobacterials | WHO-ATC code J04 | Positive |
| 54 | Antivirals for systemic use | WHO-ATC code J05 | Positive |
| 55 | Immune sera and immunoglobulins | WHO-ATC code J06 | Positive |
| 56 | Vaccines | WHO-ATC code J07 | Positive |
| 57 | Antineoplastic agents | WHO-ATC code L01 | Positive |
| 58 | Endocrine therapy | WHO-ATC code L02 | Positive |
| 59 | Immunostimulants | WHO-ATC code L03 | Positive |
| 60 | Immunosuppressants | WHO-ATC code L04 | Positive |
| 61 | Antiinflammatory and antirheumatic products | WHO-ATC code M01 | Positive |
| 62 | Topical products for joint and muscular pain | WHO-ATC code M02 | Positive |
| 63 | Muscle relaxants | WHO-ATC code M03 | Positive |
| 64 | Antigout preparations | WHO-ATC code M04 | Positive |
| 65 | Drugs for treatment of bone diseases | WHO-ATC code M05 | Positive |
| 66 | Other drugs for disorders of the musculo-skeletal system | WHO-ATC code M09 | Positive |
| 67 | Anesthetics | WHO-ATC code N01 | Positive |
| 68 | Analgesics | WHO-ATC code N02 | Positive |
| 69 | Antiepileptics | WHO-ATC code N03 | Positive |
| 70 | Anti-parkinson drugs | WHO-ATC code N04 | Positive |
| 71 | Psycholeptics | WHO-ATC code N05 | Positive |
| 72 | Psychoanaleptics | WHO-ATC code N06 | Positive |
| 73 | Other nervous system drugs | WHO-ATC code N07 | Positive |
| 74 | Antiprotozoals | WHO-ATC code P01 | Positive |
| 75 | Anthelmintics | WHO-ATC code P02 | Positive |
| 76 | Ectoparasiticides, incl. scabicides, insecticides and repellents | WHO-ATC code P03 | Positive |
| 77 | Nasal preparations | WHO-ATC code R01 | Positive |
| 78 | Throat preparations | WHO-ATC code R02 | Positive |
| 79 | Drugs for obstructive airway diseases | WHO-ATC code R03 | Positive |
| 80 | Cough and cold preparations | WHO-ATC code R05 | Positive |
| 81 | Antihistamines for systemic use | WHO-ATC code R06 | Positive |
| 82 | Other respiratory system products | WHO-ATC code R07 | Positive |
| 83 | Ophthalmologicals | WHO-ATC code S01 | Positive |
| 84 | Otologicals | WHO-ATC code S02 | Positive |
| 85 | Ophthalmological and otological preparations | WHO-ATC code S03 | Positive |
| 86 | Allergens | WHO-ATC code V01 | Positive |
| 87 | All other therapeutic products | WHO-ATC code V03 | Positive |
| 88 | Diagnostic agents | WHO-ATC code V04 | Positive |
| 89 | General nutrients | WHO-ATC code V06 | Positive |
| 90 | All other non-therapeutic products | WHO-ATC code V07 | Positive |
| 91 | Contrast media | WHO-ATC code V08 | Positive |
| 92 | Diagnostic radiopharmaceuticals | WHO-ATC code V09 | Positive |
| 93 | Therapeutic radiopharmaceuticals | WHO-ATC code V10 | Positive |
| 94 | Surgical dressings | WHO-ATC code V20 | Positive |
| Health check-up results (23 variables) | | | |
| 1 | Body mass index |  | Positive |
| 2 | Body weight |  | Positive |
| 3 | Waist circumference |  | Positive |
| 4 | Systolic blood pressure |  | Positive |
| 5 | Diastolic blood pressure |  | Positive |
| 6 | Triglycerides |  | Positive |
| 7 | High-density lipoprotein cholesterol |  | Negative |
| 8 | Low-density lipoprotein cholesterol |  | Positive |
| 9 | Non-high-density lipoprotein cholesterol |  | Positive |
| 10 | Aspartate aminotransferase |  | Positive |
| 11 | Alanine aminotransferase |  | Positive |
| 12 | Gamma-glutamyl transferase |  | Positive |
| 13 | Hemoglobin A1c |  | Positive |
| 14 | Fasting blood glucose |  | Positive |
| 15 | Casual blood glucose |  | Positive |
| 16 | Urinary glucose | Graded as 1 (−), 2 (±), 3 (+), 4 (++), and 5 (+++) | Positive |
| 17 | Urinary protein | Graded as 1 (−), 2 (±), 3 (+), 4 (++), and 5 (+++) | Positive |
| 18 | Uric acid |  | Positive |
| 19 | Creatinine |  | Positive |
| 20 | Estimated glomerular filtration rate |  | Negative |
| 21 | Hematocrit |  | Positive |
| 22 | Red blood cell count |  | Positive |
| 23 | Smoking status |  | Positive |
| Personal health records in Pep Up (110 variables) | | | |
| 1 | Mean systolic blood pressure (7-day) |  | Positive |
| 2 | Mean systolic blood pressure (1-month) |  | Positive |
| 3 | Mean systolic blood pressure (3-month) |  | Positive |
| 4 | Mean diastolic blood pressure (7-day) |  | Positive |
| 5 | Mean diastolic blood pressure (1-month) |  | Positive |
| 6 | Mean diastolic blood pressure (3-month) |  | Positive |
| 7 | Mean pulse pressure (7-day) |  | Positive |
| 8 | Mean pulse pressure (1-month) |  | Positive |
| 9 | Mean pulse pressure (3-month) |  | Positive |
| 10 | Mean mean arterial pressure (7-day) |  | Positive |
| 11 | Mean mean arterial pressure (1-month) |  | Positive |
| 12 | Mean mean arterial pressure (3-month) |  | Positive |
| 13 | Mean resting heart rate (7-day) |  | Positive |
| 14 | Mean resting heart rate (1-month) |  | Positive |
| 15 | Mean resting heart rate (3-month) |  | Positive |
| 16 | Mean shock index (7-day) |  | Positive |
| 17 | Mean shock index (1-month) |  | Positive |
| 18 | Mean shock index (3-month) |  | Positive |
| 19 | Mean double product (7-day) |  | Positive |
| 20 | Mean double product (1-month) |  | Positive |
| 21 | Mean double product (3-month) |  | Positive |
| 22 | Mean morning systolic blood pressure (7-day) | Measured between 4:00 and 12:00 | Positive |
| 23 | Mean morning systolic blood pressure (1-month) | Measured between 4:00 and 12:00 | Positive |
| 24 | Mean morning systolic blood pressure (3-month) | Measured between 4:00 and 12:00 | Positive |
| 25 | Mean morning diastolic blood pressure (7-day) | Measured between 4:00 and 12:00 | Positive |
| 26 | Mean morning diastolic blood pressure (1-month) | Measured between 4:00 and 12:00 | Positive |
| 27 | Mean morning diastolic blood pressure (3-month) | Measured between 4:00 and 12:00 | Positive |
| 28 | Mean evening systolic blood pressure (7-day) | Measured between 18:00 and 2:00 | Positive |
| 29 | Mean evening systolic blood pressure (1-month) | Measured between 18:00 and 2:00 | Positive |
| 30 | Mean evening systolic blood pressure (3-month) | Measured between 18:00 and 2:00 | Positive |
| 31 | Mean evening diastolic blood pressure (7-day) | Measured between 18:00 and 2:00 | Positive |
| 32 | Mean evening diastolic blood pressure (1-month) | Measured between 18:00 and 2:00 | Positive |
| 33 | Mean evening diastolic blood pressure (3-month) | Measured between 18:00 and 2:00 | Positive |
| 34 | Mean excess systolic blood pressure above the reference level (7-day) | The reference level is defined as 129 mmHg | Positive |
| 35 | Mean excess systolic blood pressure above the reference level (1-month) | The reference level is defined as 129 mmHg | Positive |
| 36 | Mean excess systolic blood pressure above the reference level (3-month) | The reference level is defined as 129 mmHg | Positive |
| 37 | Mean excess diastolic blood pressure above the reference level (7-day) | The reference level is defined as 84 mmHg | Positive |
| 38 | Mean excess diastolic blood pressure above the reference level (1-month) | The reference level is defined as 84 mmHg | Positive |
| 39 | Mean excess diastolic blood pressure above the reference level (3-month) | The reference level is defined as 84 mmHg | Positive |
| 40 | Difference between health checkup systolic blood pressure and mean systolic blood pressure (3-month) | Health checkup systolic blood pressure minus mean systolic blood pressure (3-month) | Negative |
| 41 | Difference between health checkup diastolic blood pressure and mean diastolic blood pressure (3-month) | Health checkup diastolic blood pressure minus mean diastolic blood pressure (3-month) | Negative |
| 42 | Difference between mean morning and evening systolic blood pressure (7-day) | Morning minus evening | Positive |
| 43 | Difference between mean morning and evening systolic blood pressure (1-month) | Morning minus evening | Positive |
| 44 | Difference between mean morning and evening systolic blood pressure (3-month) | Morning minus evening | Positive |
| 45 | Difference between mean morning and evening diastolic blood pressure (7-day) | Morning minus evening | Positive |
| 46 | Difference between mean morning and evening diastolic blood pressure (1-month) | Morning minus evening | Positive |
| 47 | Difference between mean morning and evening diastolic blood pressure (3-month) | Morning minus evening | Positive |
| 48 | Change in mean systolic blood pressure (from 1-month to 7-day) |  | Positive |
| 49 | Change in mean systolic blood pressure (from 3-month to 7-day) |  | Positive |
| 50 | Change in mean systolic blood pressure (from 3-month to 1-month) |  | Positive |
| 51 | Change in mean diastolic blood pressure (from 1-month to 7-day) |  | Positive |
| 52 | Change in mean diastolic blood pressure (from 3-month to 7-day) |  | Positive |
| 53 | Change in mean diastolic blood pressure (from 3-month to 1-month) |  | Positive |
| 54 | Change in mean pulse pressure (from 1-month to 7-day) |  | Positive |
| 55 | Change in mean pulse pressure (from 3-month to 7-day) |  | Positive |
| 56 | Change in mean pulse pressure (from 3-month to 1-month) |  | Positive |
| 57 | Change in mean mean arterial pressure (from 1-month to 7-day) |  | Positive |
| 58 | Change in mean mean arterial pressure (from 3-month to 7-day) |  | Positive |
| 59 | Change in mean mean arterial pressure (from 3-month to 1-month) |  | Positive |
| 60 | Change in mean resting heart rate (from 1-month to 7-day) |  | Positive |
| 61 | Change in mean resting heart rate (from 3-month to 7-day) |  | Positive |
| 62 | Change in mean resting heart rate (from 3-month to 1-month) |  | Positive |
| 63 | Change in mean shock index (from 1-month to 7-day) |  | Positive |
| 64 | Change in mean shock index (from 3-month to 7-day) |  | Positive |
| 65 | Change in mean shock index (from 3-month to 1-month) |  | Positive |
| 66 | Change in mean double product (from 1-month to 7-day) |  | Positive |
| 67 | Change in mean double product (from 3-month to 7-day) |  | Positive |
| 68 | Change in mean double product (from 3-month to 1-month) |  | Positive |
| 69 | Change in mean morning systolic blood pressure (from 1-month to 7-day) | Measured between 4:00 and 12:00 | Positive |
| 70 | Change in mean morning systolic blood pressure (from 3-month to 7-day) | Measured between 4:00 and 12:00 | Positive |
| 71 | Change in mean morning systolic blood pressure (from 3-month to 1-month) | Measured between 4:00 and 12:00 | Positive |
| 72 | Change in mean morning diastolic blood pressure (from 1-month to 7-day) | Measured between 4:00 and 12:00 | Positive |
| 73 | Change in mean morning diastolic blood pressure (from 3-month to 7-day) | Measured between 4:00 and 12:00 | Positive |
| 74 | Change in mean morning diastolic blood pressure (from 3-month to 1-month) | Measured between 4:00 and 12:00 | Positive |
| 75 | Change in mean evening systolic blood pressure (from 1-month to 7-day) | Measured between 18:00 and 2:00 | Positive |
| 76 | Change in mean evening systolic blood pressure (from 3-month to 7-day) | Measured between 18:00 and 2:00 | Positive |
| 77 | Change in mean evening systolic blood pressure (from 3-month to 1-month) | Measured between 18:00 and 2:00 | Positive |
| 78 | Change in mean evening diastolic blood pressure (from 1-month to 7-day) | Measured between 18:00 and 2:00 | Positive |
| 79 | Change in mean evening diastolic blood pressure (from 3-month to 7-day) | Measured between 18:00 and 2:00 | Positive |
| 80 | Change in mean evening diastolic blood pressure (from 3-month to 1-month) | Measured between 18:00 and 2:00 | Positive |
| 81 | Change in mean excess systolic blood pressure above the reference level (from 1-month to 7-day) | The reference level is defined as 129 mmHg | Positive |
| 82 | Change in mean excess systolic blood pressure above the reference level (from 3-month to 7-day) | The reference level is defined as 129 mmHg | Positive |
| 83 | Change in mean excess systolic blood pressure above the reference level (from 3-month to 1-month) | The reference level is defined as 129 mmHg | Positive |
| 84 | Change in mean excess diastolic blood pressure above the reference level (from 1-month to 7-day) | The reference level is defined as 84 mmHg | Positive |
| 85 | Change in mean excess diastolic blood pressure above the reference level (from 3-month to 7-day) | The reference level is defined as 84 mmHg | Positive |
| 86 | Change in mean excess diastolic blood pressure above the reference level (from 3-month to 1-month) | The reference level is defined as 84 mmHg | Positive |
| 87 | Mean body weight (7-day) |  | Positive |
| 88 | Mean body weight (1-month) |  | Positive |
| 89 | Mean body weight (3-month) |  | Positive |
| 90 | Mean body fat percentage (7-day) |  | Positive |
| 91 | Mean body fat percentage (1-month) |  | Positive |
| 92 | Mean body fat percentage (3-month) |  | Positive |
| 93 | Mean steps (7-day) |  | Negative |
| 94 | Mean steps (1-month) |  | Negative |
| 95 | Mean steps (3-month) |  | Negative |
| 96 | Mean sleep duration (7-day) |  | Negative |
| 97 | Mean sleep duration (1-month) |  | Negative |
| 98 | Mean sleep duration (3-month) |  | Negative |
| 99 | Change in mean body weight (from 1-month to 7-day) |  | Positive |
| 100 | Change in mean body weight (from 3-month to 7-day) |  | Positive |
| 101 | Change in mean body weight (from 3-month to 1-month) |  | Positive |
| 102 | Change in mean body fat percentage (from 1-month to 7-day) |  | Positive |
| 103 | Change in mean body fat percentage (from 3-month to 7-day) |  | Positive |
| 104 | Change in mean body fat percentage (from 3-month to 1-month) |  | Positive |
| 105 | Change in mean steps (from 1-month to 7-day) |  | Negative |
| 106 | Change in mean steps (from 3-month to 7-day) |  | Negative |
| 107 | Change in mean steps (from 3-month to 1-month) |  | Negative |
| 108 | Change in mean sleep duration (from 1-month to 7-day) |  | Negative |
| 109 | Change in mean sleep duration (from 3-month to 7-day) |  | Negative |
| 110 | Change in mean sleep duration (from 3-month to 1-month) |  | Negative |
