## Supplementary material for "Development and internal validation of a prediction model for sleep apnea syndrome treated with continuous positive airway pressure based on claims and health checkup data linked to personal health records": Online Resource 2

**Online Resource 2. Distribution of predictor variables**

|  | Variable name | Number (%) of missing data  (N = 18,692,873) | Distribution (mean±SD for continuous variables, number (%) for binary or categorical variables) |
| --- | --- | --- | --- |
| Basic information (2 variables) | | | |
| 1 | Sex | 0 (0.0%) | Men: 11,682,035 (62.5)  Women: 7,010,838 (37.5) |
| 2 | Age | 0 (0.0%) | 44.8 ± 11.3 |
| Diagnoses in the claims data (50 variables) | | | |
| 1 | Cancer | 0 (0.0%) | 0.11 ± 0.86 |
| 2 | Polyp | 0 (0.0%) | 0.16 ± 0.95 |
| 3 | Myocardial infarction | 0 (0.0%) | 0.01 ± 0.25 |
| 4 | Angina pectoris | 0 (0.0%) | 0.10 ± 0.87 |
| 5 | Hypertension | 0 (0.0%) | 0.93 ± 2.65 |
| 6 | Arrhythmia | 0 (0.0%) | 0.12 ± 0.96 |
| 7 | Stroke | 0 (0.0%) | 0.04 ± 0.51 |
| 8 | Dementia | 0 (0.0%) | 0.00 ± 0.19 |
| 9 | Schizophrenia | 0 (0.0%) | 0.07 ± 0.80 |
| 10 | Depressive disorder | 0 (0.0%) | 0.31 ± 1.67 |
| 11 | Neurotic disorder | 0 (0.0%) | 0.31 ± 1.59 |
| 12 | Eating disorder | 0 (0.0%) | 0.00 ± 0.13 |
| 13 | Developmental disorder | 0 (0.0%) | 0.03 ± 0.55 |
| 14 | Insomnia | 0 (0.0%) | 0.39 ± 1.82 |
| 15 | Autonomic nervous system disorder | 0 (0.0%) | 0.03 ± 0.47 |
| 16 | Epilepsy | 0 (0.0%) | 0.04 ± 0.58 |
| 17 | Asthma | 0 (0.0%) | 0.31 ± 1.39 |
| 18 | Tuberculosis | 0 (0.0%) | 0.00 ± 0.11 |
| 19 | Chronic obstructive pulmonary disease | 0 (0.0%) | 0.01 ± 0.29 |
| 20 | Gastroesophageal reflux disease | 0 (0.0%) | 0.86 ± 2.27 |
| 21 | Gastric or duodenal ulcer | 0 (0.0%) | 0.17 ± 1.06 |
| 22 | Inflammatory bowel disease | 0 (0.0%) | 0.03 ± 0.50 |
| 23 | Hepatitis | 0 (0.0%) | 0.09 ± 0.80 |
| 24 | Liver cirrhosis | 0 (0.0%) | 0.01 ± 0.27 |
| 25 | Fatty liver disease | 0 (0.0%) | 0.31 ± 1.49 |
| 26 | Pancreatitis | 0 (0.0%) | 0.03 ± 0.40 |
| 27 | Nephritis | 0 (0.0%) | 0.03 ± 0.43 |
| 28 | Renal failure | 0 (0.0%) | 0.05 ± 0.62 |
| 29 | Urolithiasis | 0 (0.0%) | 0.05 ± 0.54 |
| 30 | Cataract | 0 (0.0%) | 0.08 ± 0.62 |
| 31 | Glaucoma | 0 (0.0%) | 0.19 ± 0.96 |
| 32 | Meniere’s disease | 0 (0.0%) | 0.04 ± 0.51 |
| 33 | Cervical dysplasia | 0 (0.0%) | 0.02 ± 0.32 |
| 34 | Uterine fibroid | 0 (0.0%) | 0.08 ± 0.64 |
| 35 | Ovarian cyst | 0 (0.0%) | 0.03 ± 0.36 |
| 36 | Mastopathy | 0 (0.0%) | 0.03 ± 0.33 |
| 37 | Miscarriage / preterm delivery | 0 (0.0%) | 0.03 ± 0.31 |
| 38 | Cesarean section | 0 (0.0%) | 0.00 ± 0.05 |
| 39 | Endometriosis | 0 (0.0%) | 0.06 ± 0.57 |
| 40 | Fracture/rupture | 0 (0.0%) | 0.11 ± 0.72 |
| 41 | Diabetes mellitus | 0 (0.0%) | 0.50 ± 1.99 |
| 42 | Thyroid disease | 0 (0.0%) | 0.14 ± 0.96 |
| 43 | Dyslipidemia | 0 (0.0%) | 0.97 ± 2.64 |
| 44 | Rheumatoid arthritis | 0 (0.0%) | 0.05 ± 0.60 |
| 45 | Osteoarthritis | 0 (0.0%) | 0.18 ± 1.14 |
| 46 | Collagen disease | 0 (0.0%) | 0.03 ± 0.47 |
| 47 | Intervertebral disc herniation | 0 (0.0%) | 0.13 ± 0.92 |
| 48 | Premenstrual syndrome | 0 (0.0%) | 0.00 ± 0.17 |
| 49 | Hyperuricemia | 0 (0.0%) | 0.33 ± 1.61 |
| 50 | Gout | 0 (0.0%) | 0.09 ± 0.80 |
| Prescriptions in the claims data (94 variables) | | | |
| 1 | Stomatological preparations | 0 (0.0%) | 0.01 ± 0.15 |
| 2 | Drugs for acid related disorders | 0 (0.0%) | 0.73 ± 1.78 |
| 3 | Drugs for functional gastrointestinal disorders | 0 (0.0%) | 0.22 ± 0.84 |
| 4 | Antiemetics and antinauseants | 0 (0.0%) | 0.01 ± 0.17 |
| 5 | Bile and liver therapy | 0 (0.0%) | 0.05 ± 0.60 |
| 6 | Drugs for constipation | 0 (0.0%) | 0.10 ± 0.58 |
| 7 | Antidiarrheals, intestinal antiinflammatory/antiinfective agents | 0 (0.0%) | 0.20 ± 0.90 |
| 8 | Antiobesity preparations, excl. diet products | 0 (0.0%) | 0.00 ± 0.03 |
| 9 | Digestives, incl. enzymes | 0 (0.0%) | 0.01 ± 0.26 |
| 10 | Drugs used in diabetes | 0 (0.0%) | 0.27 ± 1.50 |
| 11 | Vitamins | 0 (0.0%) | 0.14 ± 0.91 |
| 12 | Mineral supplements | 0 (0.0%) | 0.02 ± 0.40 |
| 13 | Tonics | 0 (0.0%) | 0.00 ± 0.00 |
| 14 | Anabolic agents for systemic use | 0 (0.0%) | 0.00 ± 0.03 |
| 15 | Appetite stimulants | 0 (0.0%) | 0.00 ± 0.01 |
| 16 | Other alimentary tract and metabolism products | 0 (0.0%) | 0.02 ± 0.30 |
| 17 | Antithrombotic agents | 0 (0.0%) | 0.12 ± 0.92 |
| 18 | Antihemorrhagics | 0 (0.0%) | 0.29 ± 0.81 |
| 19 | Antianemic preparations | 0 (0.0%) | 0.15 ± 0.86 |
| 20 | Blood substitutes and perfusion solutions | 0 (0.0%) | 0.24 ± 0.81 |
| 21 | Other hematological agents | 0 (0.0%) | 0.00 ± 0.01 |
| 22 | Cardiac therapy | 0 (0.0%) | 0.08 ± 0.60 |
| 23 | Antihypertensives | 0 (0.0%) | 0.02 ± 0.44 |
| 24 | Diuretics | 0 (0.0%) | 0.08 ± 0.75 |
| 25 | Peripheral vasodilators | 0 (0.0%) | 0.03 ± 0.40 |
| 26 | Vasoprotectives | 0 (0.0%) | 0.22 ± 0.95 |
| 27 | Beta blocking agents | 0 (0.0%) | 0.12 ± 0.97 |
| 28 | Calcium channel blockers | 0 (0.0%) | 0.43 ± 1.81 |
| 29 | Agents acting on the renin-angiotensin system | 0 (0.0%) | 0.56 ± 2.08 |
| 30 | Lipid modifying agents | 0 (0.0%) | 0.68 ± 2.24 |
| 31 | Antifungals for dermatological use | 0 (0.0%) | 0.09 ± 0.55 |
| 32 | Emollients and protectives | 0 (0.0%) | 0.09 ± 0.55 |
| 33 | Preparations for treatment of wounds and ulcers | 0 (0.0%) | 0.02 ± 0.21 |
| 34 | Antipruritics, incl. antihistamines, anesthetics, etc. | 0 (0.0%) | 0.02 ± 0.20 |
| 35 | Antipsoriatics | 0 (0.0%) | 0.01 ± 0.25 |
| 36 | Antibiotics and chemotherapeutics for dermatological use | 0 (0.0%) | 0.08 ± 0.40 |
| 37 | Corticosteroids, dermatological preparations | 0 (0.0%) | 0.37 ± 1.12 |
| 38 | Antiseptics and disinfectants | 0 (0.0%) | 0.03 ± 0.28 |
| 39 | Medicated dressings | 0 (0.0%) | 0.00 ± 0.02 |
| 40 | Anti-acne preparations | 0 (0.0%) | 0.07 ± 0.49 |
| 41 | Other dermatological preparations | 0 (0.0%) | 0.08 ± 0.61 |
| 42 | Gynecological antiinfectives and antiseptics | 0 (0.0%) | 0.02 ± 0.21 |
| 43 | Other gynecologicals | 0 (0.0%) | 0.02 ± 0.38 |
| 44 | Sex hormones and modulators of the genital system | 0 (0.0%) | 0.13 ± 0.85 |
| 45 | Urologicals | 0 (0.0%) | 0.06 ± 0.62 |
| 46 | Pituitary and hypothalamic hormones and analogues | 0 (0.0%) | 0.01 ± 0.14 |
| 47 | Corticosteroids for systemic use | 0 (0.0%) | 0.22 ± 0.92 |
| 48 | Thyroid therapy | 0 (0.0%) | 0.05 ± 0.58 |
| 49 | Pancreatic hormones | 0 (0.0%) | 0.00 ± 0.07 |
| 50 | Calcium homeostasis | 0 (0.0%) | 0.01 ± 0.23 |
| 51 | Antibacterials for systemic use | 0 (0.0%) | 0.47 ± 1.04 |
| 52 | Antimycotics for systemic use | 0 (0.0%) | 0.00 ± 0.12 |
| 53 | Antimycobacterials | 0 (0.0%) | 0.00 ± 0.09 |
| 54 | Antivirals for systemic use | 0 (0.0%) | 0.07 ± 0.37 |
| 55 | Immune sera and immunoglobulins | 0 (0.0%) | 0.00 ± 0.12 |
| 56 | Vaccines | 0 (0.0%) | 0.00 ± 0.05 |
| 57 | Antineoplastic agents | 0 (0.0%) | 0.02 ± 0.38 |
| 58 | Endocrine therapy | 0 (0.0%) | 0.03 ± 0.38 |
| 59 | Immunostimulants | 0 (0.0%) | 0.00 ± 0.09 |
| 60 | Immunosuppressants | 0 (0.0%) | 0.05 ± 0.60 |
| 61 | Antiinflammatory and antirheumatic products | 0 (0.0%) | 0.53 ± 1.20 |
| 62 | Topical products for joint and muscular pain | 0 (0.0%) | 0.31 ± 1.09 |
| 63 | Muscle relaxants | 0 (0.0%) | 0.07 ± 0.52 |
| 64 | Antigout preparations | 0 (0.0%) | 0.29 ± 1.48 |
| 65 | Drugs for treatment of bone diseases | 0 (0.0%) | 0.02 ± 0.41 |
| 66 | Other drugs for disorders of the musculo-skeletal system | 0 (0.0%) | 0.04 ± 0.51 |
| 67 | Anesthetics | 0 (0.0%) | 0.22 ± 0.74 |
| 68 | Analgesics | 0 (0.0%) | 0.50 ± 1.13 |
| 69 | Antiepileptics | 0 (0.0%) | 0.06 ± 0.71 |
| 70 | Anti-parkinson drugs | 0 (0.0%) | 0.01 ± 0.34 |
| 71 | Psycholeptics | 0 (0.0%) | 0.43 ± 1.79 |
| 72 | Psychoanaleptics | 0 (0.0%) | 0.22 ± 1.42 |
| 73 | Other nervous system drugs | 0 (0.0%) | 0.04 ± 0.39 |
| 74 | Antiprotozoals | 0 (0.0%) | 0.00 ± 0.16 |
| 75 | Anthelmintics | 0 (0.0%) | 0.00 ± 0.02 |
| 76 | Ectoparasiticides, incl. scabicides, insecticides and repellents | 0 (0.0%) | 0.00 ± 0.01 |
| 77 | Nasal preparations | 0 (0.0%) | 0.27 ± 0.99 |
| 78 | Throat preparations | 0 (0.0%) | 0.11 ± 0.43 |
| 79 | Drugs for obstructive airway diseases | 0 (0.0%) | 0.30 ± 1.17 |
| 80 | Cough and cold preparations | 0 (0.0%) | 0.44 ± 1.02 |
| 81 | Antihistamines for systemic use | 0 (0.0%) | 0.65 ± 1.61 |
| 82 | Other respiratory system products | 0 (0.0%) | 0.00 ± 0.01 |
| 83 | Ophthalmologicals | 0 (0.0%) | 0.43 ± 1.15 |
| 84 | Otologicals | 0 (0.0%) | 0.03 ± 0.30 |
| 85 | Ophthalmological and otological preparations | 0 (0.0%) | 0.16 ± 0.74 |
| 86 | Allergens | 0 (0.0%) | 0.03 ± 0.45 |
| 87 | All other therapeutic products | 0 (0.0%) | 0.40 ± 1.33 |
| 88 | Diagnostic agents | 0 (0.0%) | 0.04 ± 0.23 |
| 89 | General nutrients | 0 (0.0%) | 0.00 ± 0.15 |
| 90 | All other non-therapeutic products | 0 (0.0%) | 0.08 ± 0.53 |
| 91 | Contrast media | 0 (0.0%) | 0.05 ± 0.29 |
| 92 | Diagnostic radiopharmaceuticals | 0 (0.0%) | 0.00 ± 0.05 |
| 93 | Therapeutic radiopharmaceuticals | 0 (0.0%) | 0.00 ± 0.01 |
| 94 | Surgical dressings | 0 (0.0%) | 0.00 ± 0.00 |
| Health check-up results (23 variables) | | | |
| 1 | Body mass index | 2054943 (11.0%) | 23.24 ± 3.92 |
| 2 | Body weight | 2055096 (11.0%) | 64.99 ± 13.49 |
| 3 | Waist circumference | 2684931 (14.4%) | 82.02 ± 10.61 |
| 4 | Systolic blood pressure | 2054943 (11.0%) | 119.93 ± 15.69 |
| 5 | Diastolic blood pressure | 2054943 (11.0%) | 74.64 ± 11.68 |
| 6 | Triglycerides | 2054943 (11.0%) | 104.86 ± 80.65 |
| 7 | High-density lipoprotein cholesterol | 2054943 (11.0%) | 63.91 ± 16.78 |
| 8 | Low-density lipoprotein cholesterol | 2054943 (11.0%) | 120.86 ± 30.62 |
| 9 | Non-high-density lipoprotein cholesterol | 14651808 (78.4%) | 140.72 ± 34.43 |
| 10 | Aspartate aminotransferase | 2054943 (11.0%) | 22.89 ± 11.30 |
| 11 | Alanine aminotransferase | 2054943 (11.0%) | 24.89 ± 20.17 |
| 12 | Gamma-glutamyl transferase | 2054943 (11.0%) | 36.86 ± 43.06 |
| 13 | Hemoglobin A1c | 2805295 (15.0%) | 5.50 ± 0.55 |
| 14 | Fasting blood glucose | 4453742 (23.8%) | 94.84 ± 15.91 |
| 15 | Casual blood glucose | 17082821 (91.4%) | 93.92 ± 19.45 |
| 16 | Urinary glucose | 2058769 (11.0%) | 1.11 ± 0.61 |
| 17 | Urinary protein | 2054943 (11.0%) | 1.12 ± 0.41 |
| 18 | Uric acid | 10379452 (55.5%) | 5.59 ± 1.41 |
| 19 | Creatinine | 6458846 (34.6%) | 0.81 ± 0.24 |
| 20 | Estimated glomerular filtration rate | 6275848 (33.6%) | 78.36 ± 15.14 |
| 21 | Hematocrit | 7824026 (41.9%) | 43.79 ± 4.10 |
| 22 | Red blood cell count | 5638464 (30.2%) | 471.25 ± 67.72 |
| 23 | Smoking status | 2214790 (11.8%) | Smoker: 3,109,552 (18.9)  Non-smoker: 13,368,531 (81.1) |
| Personal health records in Pep Up (110 variables) | | | |
| 1 | Mean systolic blood pressure (7-day) | 18608027 (99.5%) | 124.52 ± 12.56 |
| 2 | Mean systolic blood pressure (1-month) | 18604788 (99.5%) | 124.68 ± 12.08 |
| 3 | Mean systolic blood pressure (3-month) | 18600671 (99.5%) | 124.56 ± 12.04 |
| 4 | Mean diastolic blood pressure (7-day) | 18608020 (99.5%) | 80.66 ± 10.02 |
| 5 | Mean diastolic blood pressure (1-month) | 18604780 (99.5%) | 80.80 ± 9.71 |
| 6 | Mean diastolic blood pressure (3-month) | 18600651 (99.5%) | 80.89 ± 9.75 |
| 7 | Mean pulse pressure (7-day) | 18608046 (99.5%) | 43.85 ± 10.01 |
| 8 | Mean pulse pressure (1-month) | 18604807 (99.5%) | 43.87 ± 9.58 |
| 9 | Mean pulse pressure (3-month) | 18600695 (99.5%) | 43.66 ± 10.28 |
| 10 | Mean mean arterial pressure (7-day) | 18608046 (99.5%) | 95.29 ± 9.85 |
| 11 | Mean mean arterial pressure (1-month) | 18604807 (99.5%) | 95.43 ± 9.54 |
| 12 | Mean mean arterial pressure (3-month) | 18600695 (99.5%) | 95.45 ± 9.38 |
| 13 | Mean resting heart rate (7-day) | 18087216 (96.8%) | 65.34 ± 9.09 |
| 14 | Mean resting heart rate (1-month) | 18071592 (96.7%) | 65.28 ± 8.74 |
| 15 | Mean resting heart rate (3-month) | 18067448 (96.7%) | 65.37 ± 8.53 |
| 16 | Mean shock index (7-day) | 18682676 (99.9%) | 0.58 ± 0.10 |
| 17 | Mean shock index (1-month) | 18682127 (99.9%) | 0.58 ± 0.09 |
| 18 | Mean shock index (3-month) | 18681810 (99.9%) | 0.57 ± 0.09 |
| 19 | Mean double product (7-day) | 18682676 (99.9%) | 8950.58 ± 1549.31 |
| 20 | Mean double product (1-month) | 18682127 (99.9%) | 8940.39 ± 1504.72 |
| 21 | Mean double product (3-month) | 18681810 (99.9%) | 8891.56 ± 1479.90 |
| 22 | Mean morning systolic blood pressure (7-day) | 18626010 (99.6%) | 125.36 ± 12.35 |
| 23 | Mean morning systolic blood pressure (1-month) | 18622838 (99.6%) | 125.54 ± 11.85 |
| 24 | Mean morning systolic blood pressure (3-month) | 18623243 (99.6%) | 125.68 ± 11.53 |
| 25 | Mean morning diastolic blood pressure (7-day) | 18626008 (99.6%) | 81.72 ± 9.82 |
| 26 | Mean morning diastolic blood pressure (1-month) | 18622830 (99.6%) | 81.85 ± 9.51 |
| 27 | Mean morning diastolic blood pressure (3-month) | 18623233 (99.6%) | 81.96 ± 9.33 |
| 28 | Mean evening systolic blood pressure (7-day) | 18668118 (99.9%) | 121.96 ± 13.15 |
| 29 | Mean evening systolic blood pressure (1-month) | 18667138 (99.9%) | 122.07 ± 12.58 |
| 30 | Mean evening systolic blood pressure (3-month) | 18664958 (99.9%) | 121.78 ± 12.64 |
| 31 | Mean evening diastolic blood pressure (7-day) | 18668113 (99.9%) | 77.70 ± 10.32 |
| 32 | Mean evening diastolic blood pressure (1-month) | 18667130 (99.9%) | 77.81 ± 10.03 |
| 33 | Mean evening diastolic blood pressure (3-month) | 18664938 (99.9%) | 78.16 ± 10.31 |
| 34 | Mean excess systolic blood pressure above the reference level (7-day) | 18608027 (99.5%) | 3.53 ± 6.02 |
| 35 | Mean excess systolic blood pressure above the reference level (1-month) | 18604788 (99.5%) | 3.59 ± 5.72 |
| 36 | Mean excess systolic blood pressure above the reference level (3-month) | 18600671 (99.5%) | 3.59 ± 5.54 |
| 37 | Mean excess diastolic blood pressure above the reference level (7-day) | 18608020 (99.5%) | 2.82 ± 5.09 |
| 38 | Mean excess diastolic blood pressure above the reference level (1-month) | 18604780 (99.5%) | 2.87 ± 4.93 |
| 39 | Mean excess diastolic blood pressure above the reference level (3-month) | 18600651 (99.5%) | 2.97 ± 5.11 |
| 40 | Difference between health checkup systolic blood pressure and mean systolic blood pressure (3-month) | 18603001 (99.5%) | 3.06 ± 15.72 |
| 41 | Difference between health checkup diastolic blood pressure and mean diastolic blood pressure (3-month) | 18602993 (99.5%) | -0.32 ± 11.14 |
| 42 | Difference between mean morning and evening systolic blood pressure (7-day) | 18681650 (99.9%) | 4.86 ± 8.99 |
| 43 | Difference between mean morning and evening systolic blood pressure (1-month) | 18680537 (99.9%) | 4.67 ± 7.95 |
| 44 | Difference between mean morning and evening systolic blood pressure (3-month) | 18681205 (99.9%) | 4.81 ± 7.62 |
| 45 | Difference between mean morning and evening diastolic blood pressure (7-day) | 18681648 (99.9%) | 5.38 ± 6.73 |
| 46 | Difference between mean morning and evening diastolic blood pressure (1-month) | 18680536 (99.9%) | 5.16 ± 6.10 |
| 47 | Difference between mean morning and evening diastolic blood pressure (3-month) | 18681208 (99.9%) | 5.25 ± 5.81 |
| 48 | Change in mean systolic blood pressure (from 1-month to 7-day) | 18611429 (99.6%) | -0.09 ± 3.61 |
| 49 | Change in mean systolic blood pressure (from 3-month to 7-day) | 18616206 (99.6%) | -0.27 ± 4.99 |
| 50 | Change in mean systolic blood pressure (from 3-month to 1-month) | 18611702 (99.6%) | -0.18 ± 2.99 |
| 51 | Change in mean diastolic blood pressure (from 1-month to 7-day) | 18611416 (99.6%) | -0.10 ± 2.56 |
| 52 | Change in mean diastolic blood pressure (from 3-month to 7-day) | 18616194 (99.6%) | -0.24 ± 3.49 |
| 53 | Change in mean diastolic blood pressure (from 3-month to 1-month) | 18611692 (99.6%) | -0.15 ± 2.12 |
| 54 | Change in mean pulse pressure (from 1-month to 7-day) | 18611446 (99.6%) | 0.00 ± 2.92 |
| 55 | Change in mean pulse pressure (from 3-month to 7-day) | 18616223 (99.6%) | -0.03 ± 4.03 |
| 56 | Change in mean pulse pressure (from 3-month to 1-month) | 18611718 (99.6%) | -0.04 ± 2.48 |
| 57 | Change in mean mean arterial pressure (from 1-month to 7-day) | 18611446 (99.6%) | -0.09 ± 2.60 |
| 58 | Change in mean mean arterial pressure (from 3-month to 7-day) | 18616223 (99.6%) | -0.25 ± 3.57 |
| 59 | Change in mean mean arterial pressure (from 3-month to 1-month) | 18611718 (99.6%) | -0.16 ± 2.14 |
| 60 | Change in mean resting heart rate (from 1-month to 7-day) | 18110637 (96.9%) | 0.12 ± 2.56 |
| 61 | Change in mean resting heart rate (from 3-month to 7-day) | 18142528 (97.1%) | 0.06 ± 3.30 |
| 62 | Change in mean resting heart rate (from 3-month to 1-month) | 18115396 (96.9%) | -0.04 ± 1.98 |
| 63 | Change in mean shock index (from 1-month to 7-day) | 18683281 (99.9%) | 0.00 ± 0.03 |
| 64 | Change in mean shock index (from 3-month to 7-day) | 18683958 (100.0%) | 0.00 ± 0.04 |
| 65 | Change in mean shock index (from 3-month to 1-month) | 18683211 (99.9%) | 0.00 ± 0.02 |
| 66 | Change in mean double product (from 1-month to 7-day) | 18683281 (99.9%) | 13.08 ± 429.60 |
| 67 | Change in mean double product (from 3-month to 7-day) | 18683958 (100.0%) | 10.81 ± 547.38 |
| 68 | Change in mean double product (from 3-month to 1-month) | 18683211 (99.9%) | -3.82 ± 308.48 |
| 69 | Change in mean morning systolic blood pressure (from 1-month to 7-day) | 18629282 (99.7%) | -0.09 ± 3.63 |
| 70 | Change in mean morning systolic blood pressure (from 3-month to 7-day) | 18633312 (99.7%) | -0.28 ± 4.94 |
| 71 | Change in mean morning systolic blood pressure (from 3-month to 1-month) | 18628949 (99.7%) | -0.20 ± 2.93 |
| 72 | Change in mean morning diastolic blood pressure (from 1-month to 7-day) | 18629277 (99.7%) | -0.10 ± 2.51 |
| 73 | Change in mean morning diastolic blood pressure (from 3-month to 7-day) | 18633306 (99.7%) | -0.24 ± 3.37 |
| 74 | Change in mean morning diastolic blood pressure (from 3-month to 1-month) | 18628937 (99.7%) | -0.14 ± 2.01 |
| 75 | Change in mean evening systolic blood pressure (from 1-month to 7-day) | 18670452 (99.9%) | -0.14 ± 3.85 |
| 76 | Change in mean evening systolic blood pressure (from 3-month to 7-day) | 18672514 (99.9%) | -0.35 ± 5.21 |
| 77 | Change in mean evening systolic blood pressure (from 3-month to 1-month) | 18670501 (99.9%) | -0.23 ± 3.15 |
| 78 | Change in mean evening diastolic blood pressure (from 1-month to 7-day) | 18670447 (99.9%) | -0.14 ± 2.85 |
| 79 | Change in mean evening diastolic blood pressure (from 3-month to 7-day) | 18672509 (99.9%) | -0.32 ± 3.83 |
| 80 | Change in mean evening diastolic blood pressure (from 3-month to 1-month) | 18670491 (99.9%) | -0.21 ± 2.35 |
| 81 | Change in mean excess systolic blood pressure above the reference level (from 1-month to 7-day) | 18611429 (99.6%) | -0.04 ± 2.10 |
| 82 | Change in mean excess systolic blood pressure above the reference level (from 3-month to 7-day) | 18616206 (99.6%) | -0.11 ± 2.86 |
| 83 | Change in mean excess systolic blood pressure above the reference level (from 3-month to 1-month) | 18611702 (99.6%) | -0.08 ± 1.67 |
| 84 | Change in mean excess diastolic blood pressure above the reference level (from 1-month to 7-day) | 18611416 (99.6%) | -0.04 ± 1.46 |
| 85 | Change in mean excess diastolic blood pressure above the reference level (from 3-month to 7-day) | 18616194 (99.6%) | -0.10 ± 2.02 |
| 86 | Change in mean excess diastolic blood pressure above the reference level (from 3-month to 1-month) | 18611692 (99.6%) | -0.07 ± 1.25 |
| 87 | Mean body weight (7-day) | 18512086 (99.0%) | 66.77 ± 12.11 |
| 88 | Mean body weight (1-month) | 18505106 (99.0%) | 66.83 ± 12.09 |
| 89 | Mean body weight (3-month) | 18508219 (99.0%) | 66.83 ± 12.07 |
| 90 | Mean body fat percentage (7-day) | 18589182 (99.4%) | 23.38 ± 6.66 |
| 91 | Mean body fat percentage (1-month) | 18584195 (99.4%) | 23.41 ± 6.65 |
| 92 | Mean body fat percentage (3-month) | 18587059 (99.4%) | 23.40 ± 6.61 |
| 93 | Mean steps (7-day) | 16675361 (89.2%) | 7511.81 ± 3965.27 |
| 94 | Mean steps (1-month) | 16636797 (89.0%) | 7491.34 ± 3714.86 |
| 95 | Mean steps (3-month) | 16588832 (88.7%) | 7486.89 ± 3607.43 |
| 96 | Mean sleep duration (7-day) | 18287810 (97.8%) | 355.84 ± 69.88 |
| 97 | Mean sleep duration (1-month) | 18275740 (97.8%) | 356.98 ± 63.25 |
| 98 | Mean sleep duration (3-month) | 18273569 (97.8%) | 358.58 ± 60.73 |
| 99 | Change in mean body weight (from 1-month to 7-day) | 18526478 (99.1%) | -0.01 ± 0.50 |
| 100 | Change in mean body weight (from 3-month to 7-day) | 18540274 (99.2%) | -0.09 ± 0.85 |
| 101 | Change in mean body weight (from 3-month to 1-month) | 18528037 (99.1%) | -0.09 ± 0.56 |
| 102 | Change in mean body fat percentage (from 1-month to 7-day) | 18596992 (99.5%) | -0.02 ± 0.49 |
| 103 | Change in mean body fat percentage (from 3-month to 7-day) | 18605871 (99.5%) | -0.07 ± 0.76 |
| 104 | Change in mean body fat percentage (from 3-month to 1-month) | 18597927 (99.5%) | -0.05 ± 0.49 |
| 105 | Change in mean steps (from 1-month to 7-day) | 16723806 (89.5%) | -1.56 ± 1501.21 |
| 106 | Change in mean steps (from 3-month to 7-day) | 16825847 (90.0%) | -24.35 ± 1882.27 |
| 107 | Change in mean steps (from 3-month to 1-month) | 16752719 (89.6%) | -22.25 ± 1004.32 |
| 108 | Change in mean sleep duration (from 1-month to 7-day) | 18305161 (97.9%) | -1.11 ± 29.78 |
| 109 | Change in mean sleep duration (from 3-month to 7-day) | 18326766 (98.0%) | -2.59 ± 34.93 |
| 110 | Change in mean sleep duration (from 3-month to 1-month) | 18306501 (97.9%) | -1.50 ± 16.78 |
