## Supplementary material for "Development and internal validation of a prediction model for sleep apnea syndrome treated with continuous positive airway pressure based on claims and health checkup data linked to personal health records": Online Resource 3

**Online Resource 3. SHapley Additive exPlanations (SHAP) plot for the top 20 predictor variables in the prediction model for sleep apnea syndrome treated with continuous positive airway pressure in a subgroup analysis by sex**

**Men**

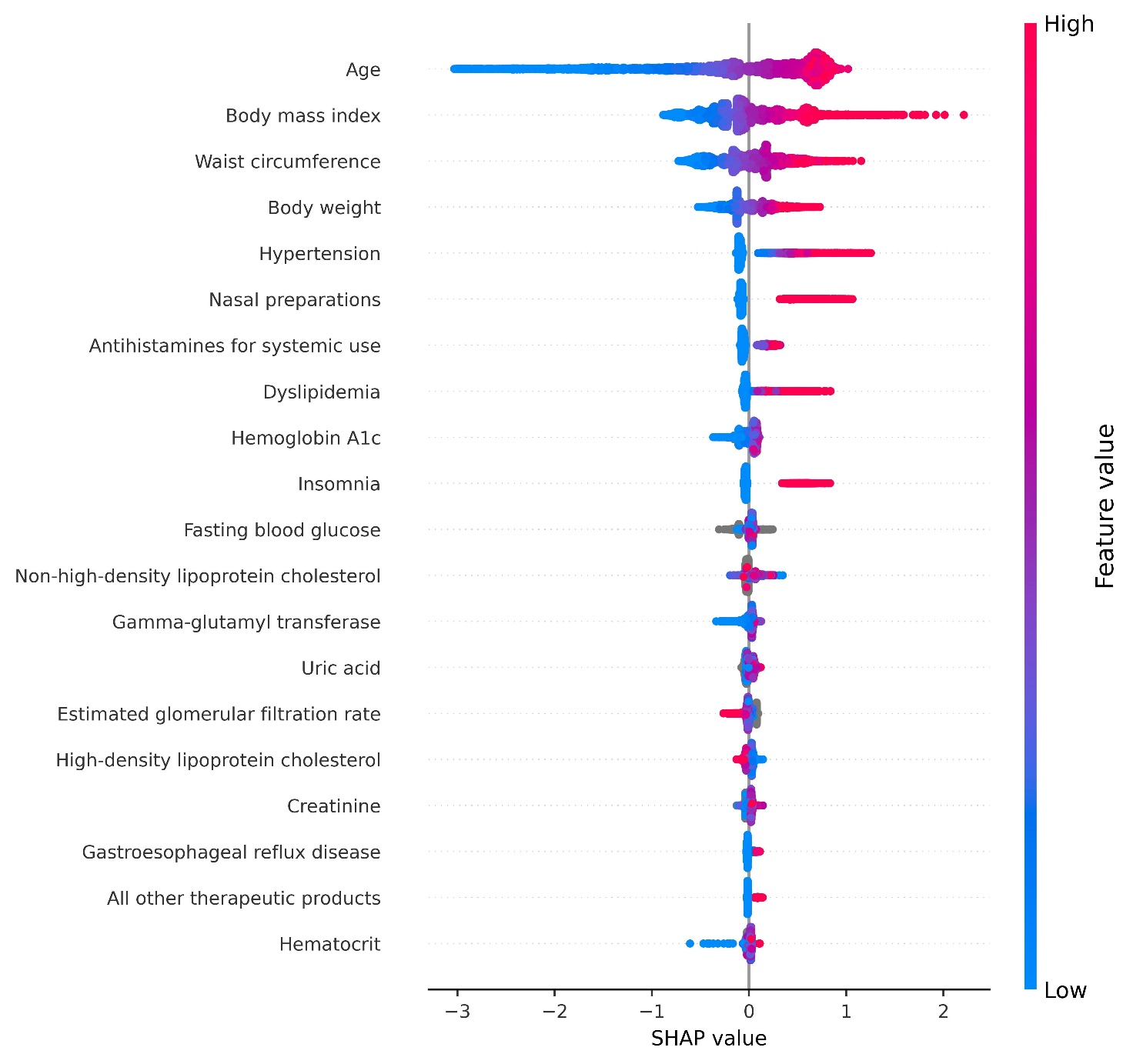

**Women**

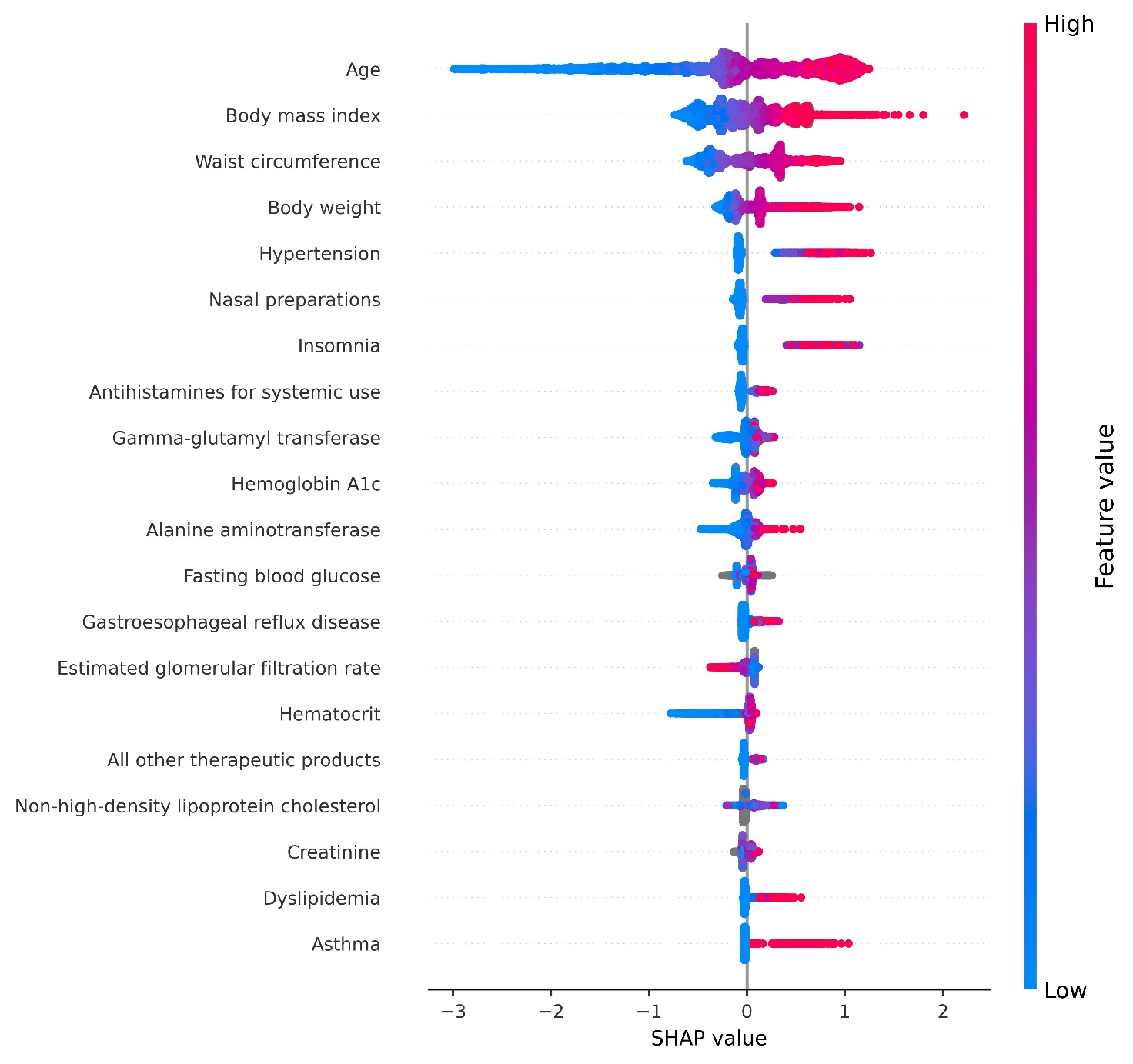
