## Supplementary material for "Development and internal validation of a prediction model for sleep apnea syndrome treated with continuous positive airway pressure based on claims and health checkup data linked to personal health records": Online Resource 4

**Online Resource 4. The area under the receiver operating characteristic curve of the prediction model for sleep apnea syndrome treated with continuous positive airway pressure in the separated validation datasets at 11 timepoints**

| Index date | AUROC (95% CI) |
| --- | --- |
| January 1, 2022 | 0.896 (0.892–0.900) |
| April 1, 2022 | 0.898 (0.895–0.902) |
| July 1, 2022 | 0.898 (0.894–0.902) |
| October 1, 2022 | 0.897 (0.894–0.901) |
| January 1, 2023 | 0.898 (0.894–0.901) |
| April 1, 2023 | 0.899 (0.895–0.902) |
| July 1, 2023 | 0.899 (0.896–0.903) |
| October 1, 2023 | 0.898 (0.894–0.901) |
| January 1, 2024 | 0.897 (0.893–0.900) |
| April 1, 2024 | 0.898 (0.895–0.902) |
| July 1, 2024 | 0.896 (0.893–0.899) |

AUROC, area under the receiver operating characteristic curve; CI, confidence interval.
